# Date of introduction and epidemiologic patterns of SARS-CoV-2 in Mogadishu, Somalia: estimates from transmission modelling of 2020 excess mortality data

**DOI:** 10.1101/2021.06.15.21258924

**Authors:** Mihaly Koltai, Abdihamid Warsame, Farah Bashiir, Terri Freemantle, Chris Williams, Mark Jit, Stefan Flasche, Nicholas G. Davies, CMMID COVID-19 working group, Ahmed Aweis, Mohamed Ahmed, Abdirisak Dalmar, Francesco Checchi

## Abstract

**Introduction:** In countries with weak surveillance systems confirmed COVID-19 deaths are likely to underestimate the death toll of the pandemic. Many countries also have incomplete vital registration systems, hampering excess mortality estimation. Here, we fitted a dynamic transmission model to satellite imagery data on burial patterns in Mogadishu, Somalia during 2020 to estimate the date of introduction, transmissibility and other epidemiologic characteristics of SARS-CoV-2 in this low-income, crisis-affected setting.

**Methods:** We performed Markov chain Monte Carlo (MCMC) fitting with an age-structured compartmental COVID-19 model to provide median estimates and credible intervals for the date of introduction, the basic reproduction number (*R*_*0*_) and the effect of non-pharmaceutical interventions in Mogadishu up to September 2020.

**Results:** Under the assumption that excess deaths in Mogadishu February-September 2020 were directly attributable to SARS-CoV-2 infection we arrived at median estimates of October-November 2019 for the date of introduction and low *R*_*0*_ estimates (1.3-1.5) stemming from the early and slow rise of excess deaths. The effect of control measures on transmissibility appeared small.

**Conclusion:** Subject to study assumptions, a very early SARS-CoV-2 introduction event may have occurred in Somalia. Estimated transmissibility in the first epidemic wave was lower than observed in European settings.

## Introduction

By May 2021, more than 3.5 million people were confirmed to have died from the COVID-19 pandemic caused by the novel SARS-CoV-2 coronavirus. While the cumulative rate of confirmed deaths has exceeded 1 per 1000 persons in the United States and many countries in Europe and Latin America, it has remained one or even two orders of magnitude lower in most of Africa(1).

While some of this difference can be potentially explained by a lower infection fatality ratio (IFR) for the entire population due to a lower median age (2–4), evidence suggests that at least critically ill COVID-19 African patients experience higher, not lower mortality than elsewhere (5), as plausibly expected due to weaker health infrastructure (6). News reports (7), studies using seroprevalence (8,9), PCR testing in morgues (10), as well as indirect data sources such as obituaries on social media (11) point to substantial under-ascertainment of cases and deaths in low-income countries, potentially ten-fold (suggested by excess mortality data from Egypt (12)) or even nearly hundred-fold (13) in crisis-ridden regions. While in high income countries (12) confirmed COVID-19 deaths are approximately in line with excess death statistics, in many African countries there are no reliable mortality statistics, precluding the use of excess deaths to infer the true scale of the pandemic.

Additionally, while the first COVID-19 cases in sub-Saharan African countries were recognised in late February (14) (16th of March in Somalia), there is considerable uncertainty about the true date of introduction, often estimated to be in January 2020 for Western Europe (15), or as early as December 2019 according to retrospective PCR on routine patient samples (16). For these reasons, alternative data sources such as the number of obituaries(11) and satellite imagery(17) of cemeteries have been leveraged to estimate the true scale of COVID-19 mortality and its early spread in African and other low- and middle-income countries.

In this study we used a dynamic transmission model to analyse a time series of excess deaths in Mogadishu (Somalia) inferred from satellite images of the six main cemeteries in the city (18). Our aim was to estimate the probable date of introduction of SARS-CoV-2, as well as the basic reproduction number (*R*_*0*_) and the effect of non-pharmaceutical interventions.

## Methods

### Data sources and statistical analysis

Details of the method for inferring excess mortality from satellite images are described in an accompanying article (18). Briefly, cemeteries in the Banadir administrative region, which contains Mogadishu, were identified via open source location data and satellite imagery (OpenStreetMap, Google Earth, GoogleMaps) by a combination of automatic image recognition and manual annotation, in addition to key informant interviews and field visits to identified cemeteries. We identified and analysed six cemeteries (Barakaat 1 and 2, Calamada, Iskool Bolisii, Kahda, Moallim Nuur). We excluded from the analysis five smaller private and family-owned cemeteries estimated to account for less than 20% of all burials because of lack of images and vegetation cover. One cemetery (Calamada) included in the analysis falls outside of Banadir region limits, but largely caters to Mogadishu residents and was therefore included.

Sixty-eight archive satellite images from the period February 2017 - September 2020 were selected on the criteria that they were cloud-free, of high radiometric quality and with a spatial resolution of 30-40 cm per pixel, were analysed through manual and semi-automated image processing to extract surface area and number of graves. An exhaustive grave count by either of these two methods was possible for 40 out of 68 satellite (58.8%) images. For the remaining images, the number of graves was extrapolated from visible areas or imputed through a generalised additive mixed model of the association between graves and surface area. Results for each image were then interpolated and summed across all cemeteries to yield a single time series of burials for the city.

To compute the baseline (pre-pandemic) crude death rate (CDR), population denominators for Mogadishu (Banadir region) were estimated using the WorldPop project’s database (19), using either the 2015 or 2019 estimates, while also adjusting for in- and out-displacement to/from the city (20). The two alternative base estimates correspond to a ‘high’ and a ‘low’ scenario with nearly identical trends (SI Figure 1) and marginally different levels (0.04-0.05 deaths/10.000 person-days) of baseline (i.e. pre-pandemic) CDR.

This level is significantly lower than previous CDR estimates for Somalia (21) between 0.2-0.6/10.000 person-days. Assuming that the level of under-estimation remains constant, we can scale the crude death rate estimated from our time series up to previous estimates (using the lower end of the estimates from (21), 0.1-0.4 deaths per 10.000 person-days). In terms of modelling transmission dynamics, such scaling of deaths merely shifts the IFR, while leaving other parameter estimates largely unchanged (SI Figure 14), and hence provides little additional information. We therefore used the observed time series of burials directly for model fitting, without scaling. To isolate excess mortality (which we assumed to be entirely attributable to SARS-CoV-2 infection: see Discussion), we extrapolated pre-2020 burial rates into the pandemic period and subtracted this baseline from the total (SI Figure 1). For excess burials (mortality) we then took the daily number of burials in the dataset and subtracted the mean level of daily burials in the four months period 01/07/2019-01/11/2019 (9.3 burials/day). We chose this pre-pandemic period as a basis of comparison as burial rates in the preceding period had been likely affected by the drought-triggered crisis 2017-2018 (21). The model output of incident deaths were fitted to this baseline-subtracted number of burials per day.

### Transmission model

We used CovidM, an age-stratified dynamic transmission model initially developed to model the spread of COVID-19 and the effect of non-pharmaceutical interventions in the UK (22,23). The model has a susceptible-exposed-infectious-recovered structure with individuals stratified into 5-year age bands. When susceptible individuals are infected they move into an exposed (incubating) compartment (E), becoming either infectious with symptoms (I_c_) following a pre-symptomatic phase (I_p_) or remaining asymptomatic (I_s_) with a lower level of infectiousness (set to 50% as in previous studies (22,24)). We used existing age-dependent estimates (22) for the proportion of individuals who are symptomatically infected (clinical fraction), as well as for the susceptibility to infection (SI Figure 7). Both of these estimates are age-dependent, with the clinical fraction 29% in the 0-9y age group and 69% above 70 years, and susceptibility among individuals aged 0-19y half of that among adults. Deaths occur in the model with a gamma-distributed (shape=22, scale=1) delay of 22 days (25,26) following the transition from exposed (E) to pre-symptomatic (I_p_) state. Other parameters of disease progression were fixed to consensus estimates in the literature (see SI Table 1). The model was parameterised with the demographic structure and contact patterns of Somalia (19), and the total population fixed to that of Mogadishu (2.2 million as of mid-2020).

### Estimates on infection fatality ratio

We used existing age-specific IFR estimates (24) demonstrating a log-linear relationship between age and the IFR. To account for the uncertainty in the IFR, we fitted the data both with the original IFR estimates from high-income countries and with upwardly adjusted IFR estimates to reflect the effect of a weak public health infrastructure. For the latter, we calculated the logit of the original IFR at each age group and increased its value (SI Figure 8), raising the mean IFR for those 75 or older from the original 11.6% to 26-70%. A possible upward shift of IFR values by age groups is supported by recent findings of substantially higher in-hospital mortality in several African countries (5). Due to its young population, the population-average IFR (calculated for a randomly chosen infected person) for Somalia would still be lower than in most high-income countries, being 0.15% with the original estimates, and population-average IFRs of 0.36%, 0.79%, 1.13% and 1.6%, respectively, under the adjusted values.

### Fitting parameters and input parameters

We estimated three epidemiological parameters of the model: the date of introduction into Mogadishu, the basic reproduction number (*R*_*0*_) and a scaling factor (*NPI_scale*) that converts the nominal stringency of non-pharmaceutical interventions (NPIs) into a relative reduction of transmissibility. We held two other parameters, the IFR and the size of the initial seeding event (defined as the number of infected individuals in compartment E), fixed and re-ran the fitting process for a range of values. Fitting all five parameters simultaneously results in strong correlations for the parameter pairs NPI_scale and IFR, *R*_*0*_ and IFR and *R*_*0*_ and seed size (SI Figure 6), suggesting they are not separately identifiable. Similarly, the seed size and the date of introduction are inversely related parameters, therefore we fitted only the date of introduction for a range of different seed sizes between 20 and 500. For simplicity we placed the seeding event on a single day; in our deterministic modelling framework a more gradual introduction does not have a significantly different effect. Initial importations were assumed to be adults between 30 and 70, an importation of younger adults leads to somewhat lower *R*_*0*_ estimates, but otherwise similar results SI Figure 15).

The effect of NPIs was accounted for by using the Oxford COVID-19 Government Response Tracker (OxCGRT) (27), using the *StringencyIndex* variable for the strength of the non-pharmaceutical interventions (NPI). Since we have no independent data source such as mobility data on the actual effect of NPIs in Somalia, we scaled the value of the stringency index by the free parameter (*NPI_scale*) that represents the effectiveness of the intervention. We distinguished three periods in terms of NPIs (SI Figure 3). The value of *StringencyIndex* increased abruptly (in three days) from 0 to 41% of its maximum at the 18th of March and stayed above 50% until the 30th of June. From the 1st of July to the 29th of August a number of relaxations followed. From the 30th of August the *StringencyIndex* started to increase again and did not decrease until the end of our fitting period. To minimise over-parameterisation, instead of using the full time series of *StringencyIndex* that could introduce significant additional complexity to model dynamics we only took the mean value of *StringencyIndex* in these three periods (0.59, 0.26 and 0.41; SI Figure 3), and implemented the effect of NPIs by reducing transmission coefficients for all age groups by the product of stringency (mean value per period) and the scaling factor (*NPI_scale*). For example, if *NPI_scale*=0.5, then in the first period when stringency was equal to 0.59 the reduction in transmission is proportional to *StringencyIndex*\**NPI_scale*=0.59*0.5, ie. 29.5%, whereas in the second period the reduction is 13% (*StringencyIndex*\**NPI_scale*=0.26*0.5).

### Time window of fitting

All fits presented in the main text were done with the time window 23/02/2020 to 24/08/2020, excluding the first smaller spike of deaths in January as well as the late spike in September. We removed the early January spike in excess burials to avoid any confounding from the two continuous weeks of reported cholera deaths in Banadir (28) that coincided with this period. Moreover this early spike of deaths is in general inconsistent with a gradually rising epidemic curve from late February. Including the deaths in late January leads to even earlier estimates for the date of introduction, but poorer fits (SI Figure 10), as the epidemiological model cannot capture this early non-monotonic dynamics.

### Fitting procedure

To estimate the unknown parameters, we fitted the CovidM model to the excess deaths time series using a Monte Carlo Markov chain (MCMC) algorithm, minimising the log-likelihood of incident deaths (assumed to be Poisson-distributed). We introduced informative prior assumptions for the date of introduction (normal distribution with mean: 01/03/2020, standard deviation: 20 days) and *R*_*0*_ (truncated normal distribution with mean=3, SD=1, bounded at 1 and 5), and an uninformative uniform distribution for the NPI scaling factor (U(0,1)). We used a differential evolution MCMC algorithm with 10 chains, with a burn-in of at least 500 iterations followed by at least 2000 samples.

## Results

Satellite imagery of the six main cemeteries in Mogadishu showed a first spike in the number of burials in late January, followed by a more sustained rise from late February (Figure 1). The weekly number of excess burials rose to approximately 60 in April and to a peak of 85 in mid-June, falling back to values near zero only in August.

**Figure 1.**
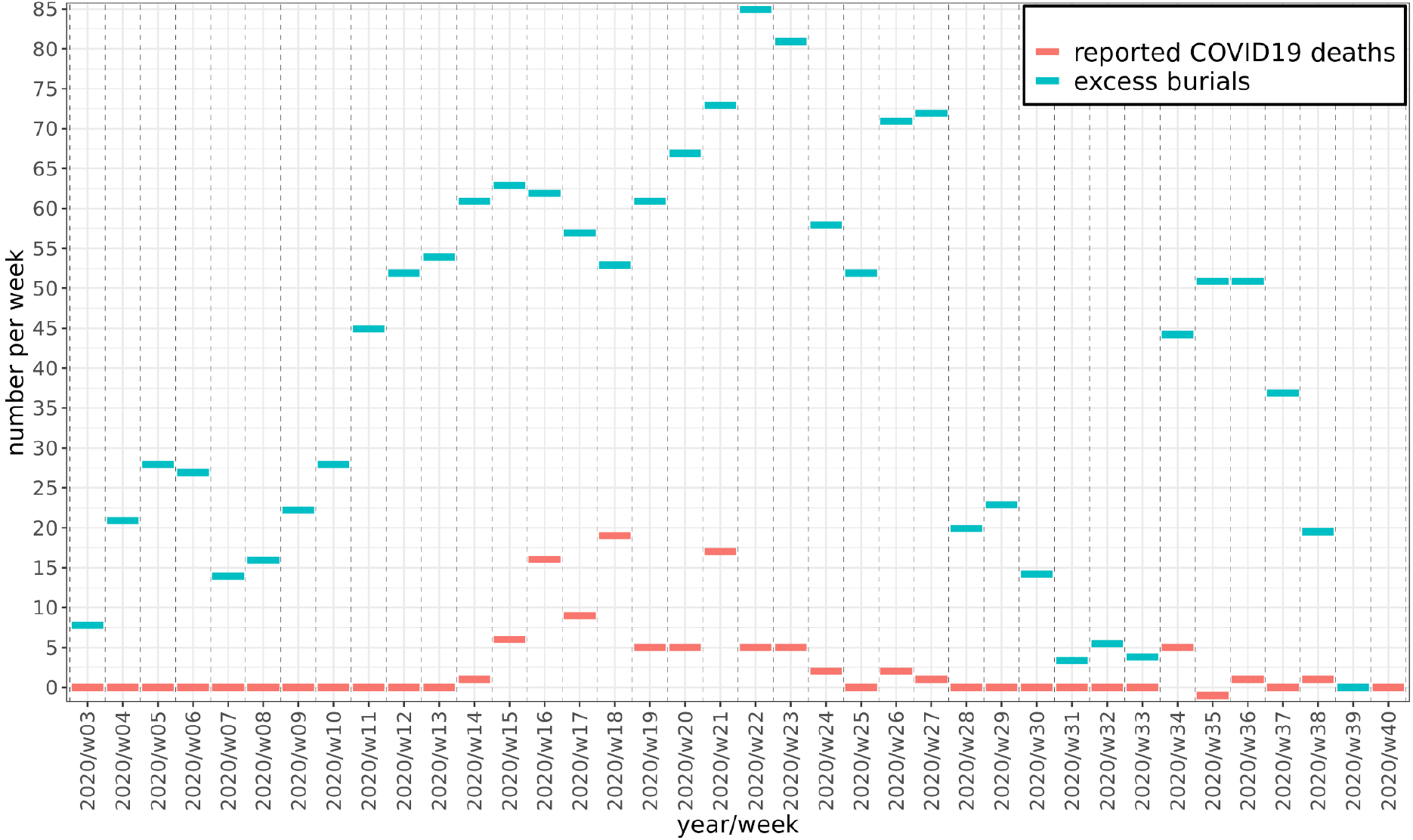
Weekly burials above the pre-pandemic baseline (excess burials) in Mogadishu compared to reported COVID-19 deaths in Somalia.

### Estimates for date of introduction and initial spread (*R*_*0*_)

The slow rise in deaths from February to mid-June and the long plateau lasting until late July results in *R*_*0*_ estimates substantially lower than those for Wuhan (29) and European countries (23) for the initial phase of the pandemic. Fitting our data with a range of IFR values (between 0.15% and 1.13%) and seed sizes resulted in median *R*_*0*_ estimates between 1.3 and 1.5 (Figure 2, Table 1). The best fits as expressed by DIC (deviance information criterion) values are for a population-wide IFR of 0.36% and 0.79% (Figure 4). These IFR values are above the high-income country-specific base assumption of 0.15% for Somalia and result in a median *R*_*0*_ estimate of 1.34-1.38 depending on the seed size (Table 1).

**Table 1:** mean values and 95% credible intervals for fitting parameters at different values of IFR and seed size

|  | IFR=0.15% | IFR=0.36% | IFR=0.79% | IFR=1.13% | IFR=1.6% | seed size |
| --- | --- | --- | --- | --- | --- | --- |
| date of introduction | 2019-10-12<br>(10-05, 10-18) | 2019-09-13<br>(08-30, 09-28) | 2019-09-29<br>(09-10,10-16) | 2019-10-06<br>(09-18, 10-21) | 2019-10-12<br>(09-27,10-27) | 20 |
|  | 2019-10-24<br>(10-18, 10-31) | 2019-09-21<br>(09-06, 10-05) | 2019-10-10<br>(09-22,10-24) | 2019-10-15<br>(10-01,10-30) | 2019-10-23<br>(10-06,11-10) | 50 |
|  | 2019-11-01<br>(10-26, 11-06) | 2019-09-30<br>(09-11, 10-19) | 2019-10-20<br>(10-07,11-03) | 2019-10-24<br>(10-08, 11-05) | 2019-10-31<br>(10-14,11-16) | 100 |
|  | 2019-11-10<br>(11-05, 11-14) | 2019-10-07<br>(09-19,10-21) | 2019-10-27<br>(10-12,11-09) | 2019-11-01<br>(10-19, 11-15) | 2019-11-11<br>(10-27,11-22) | 200 |
| $R_0$ | 1.48<br>(1.45, 1.5) | 1.38<br>(1.35,1.42) | 1.38<br>(1.33,1.42) | 1.37<br>(1.32,1.41) | 1.35<br>(1.32,1.4) | 20 |
|  | 1.48 (1.46,<br>1.5) | 1.37<br>(1.33, 1.4) | 1.37<br>(1.32,1.41) | 1.35<br>(1.31, 1.4) | 1.34<br>(1.3,1.4) | 50 |
|  | 1.47<br>(1.45,1.49) | 1.36<br>(1.31,1.41) | 1.35<br>(1.32, 1.4) | 1.33<br>(1.3, 1.37) | 1.32<br>(1.28,1.37) | 100 |
|  | 1.47<br>(1.45,1.49) | 1.35<br>(1.31,1.38) | 1.34<br>(1.3, 1.38) | 1.32<br>(1.28,1.36) | 1.31<br>(1.27,1.35) | 200 |
| $NPI\_scale$ | 0.006<br>(0,0.022) | 0.17<br>(0.12, 0.24) | 0.44<br>(0.4, 0.5) | 0.5<br>(0.45, 0.55) | 0.53<br>(0.49,0.58) | 20 |
|  | 0.006<br>(0, 0.031) | 0.14<br>(0.07, 0.2) | 0.43<br>(0.37, 0.48) | 0.477<br>(0.43, 0.54) | 0.51<br>(0.45,0.58) | 50 |
|  | 0.007<br>(0, 0.028) | 0.12<br>(0.03, 0.21) | 0.42<br>(0.37, 0.48) | 0.46<br>(0.41, 0.51) | 0.49<br>(0.43,0.55) | 100 |
|  | 0.005<br>(0, 0.018) | 0.1<br>(0.02, 0.18) | 0.39<br>(0.34, 0.45) | 0.44<br>(0.39, 0.5) | 0.48<br>(0.42,0.53) | 200 |

**Figure 2.**
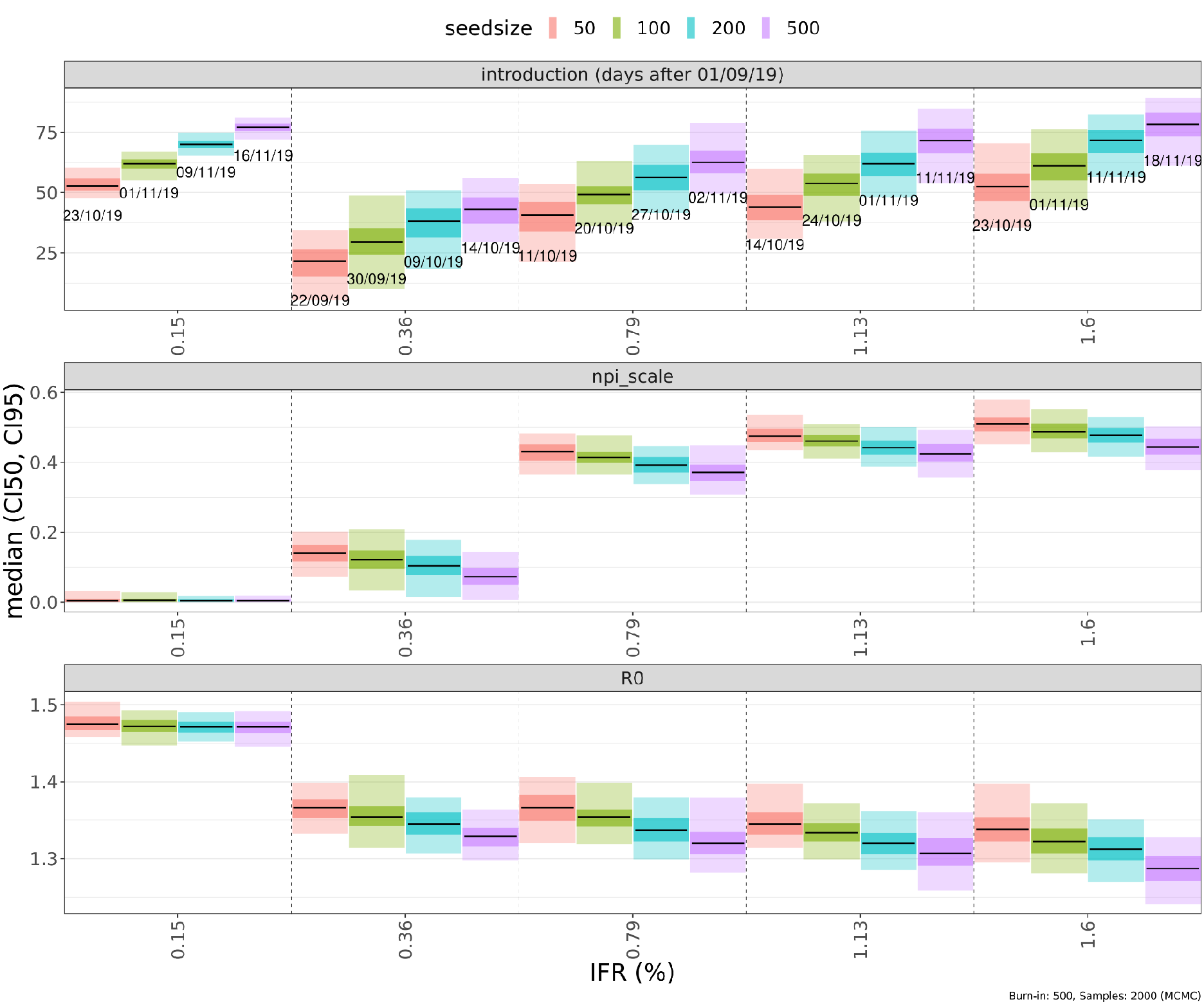
Median values and credible intervals for the fitting parameters (introduction date, *NPI_scale, R*_*0*_) and quality of fits at different assumed values of the infection fatality ratio (x-axis) and seed size (colors). In the top panel, labels below the lines show median estimates of the date of introduction. Shaded areas around the median (black) are 50% (darker) and 95% credible intervals.

**Figure 3.**
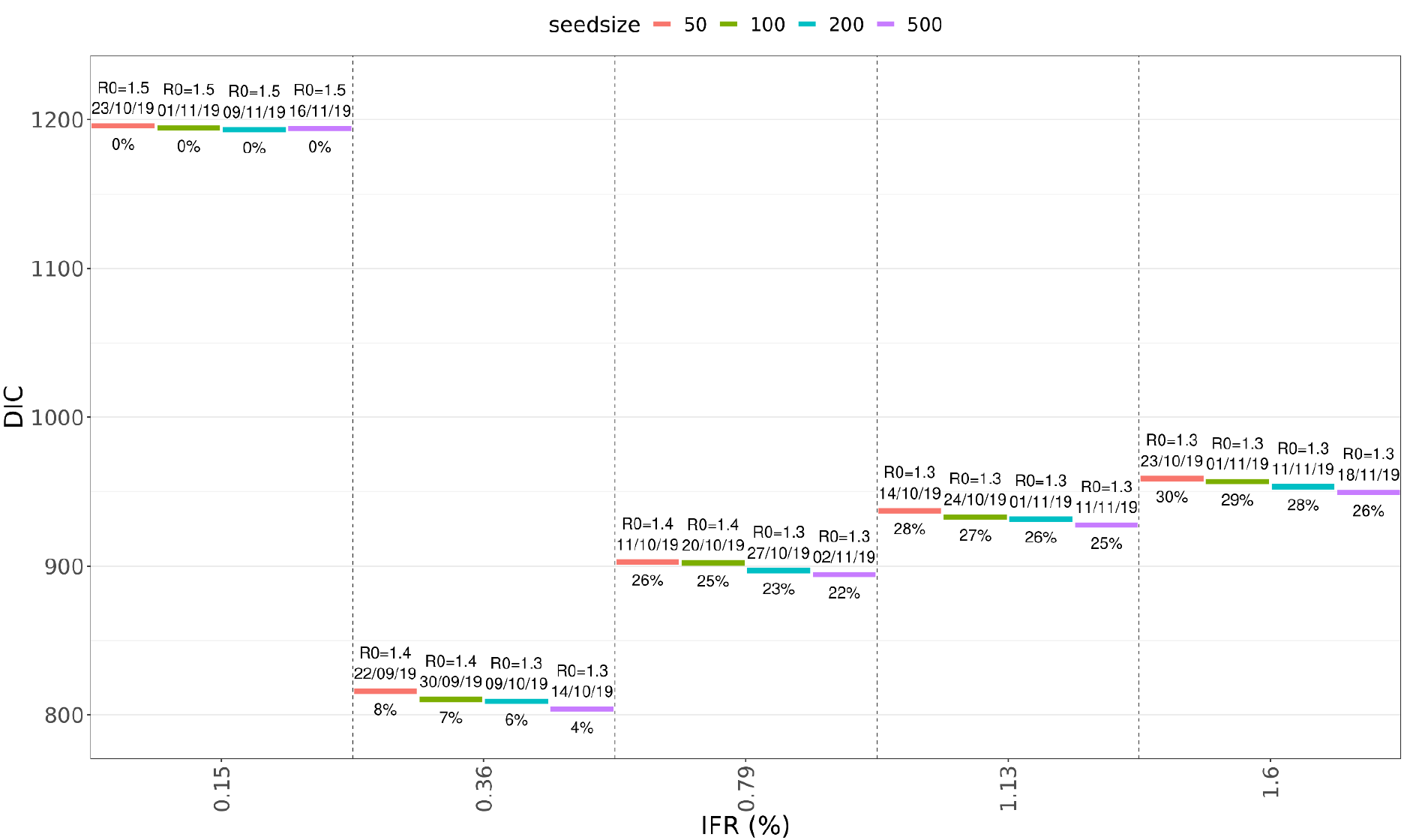
Goodness of fit as measured by DIC (deviance information criterion) at different values for seed size and population-wide IFR. The labels above the colored lines show median estimates for *R*_*0*_ and the date of introduction, and the NPI-induced reduction in transmissibility during the first NPI period below.

**Figure 4.**
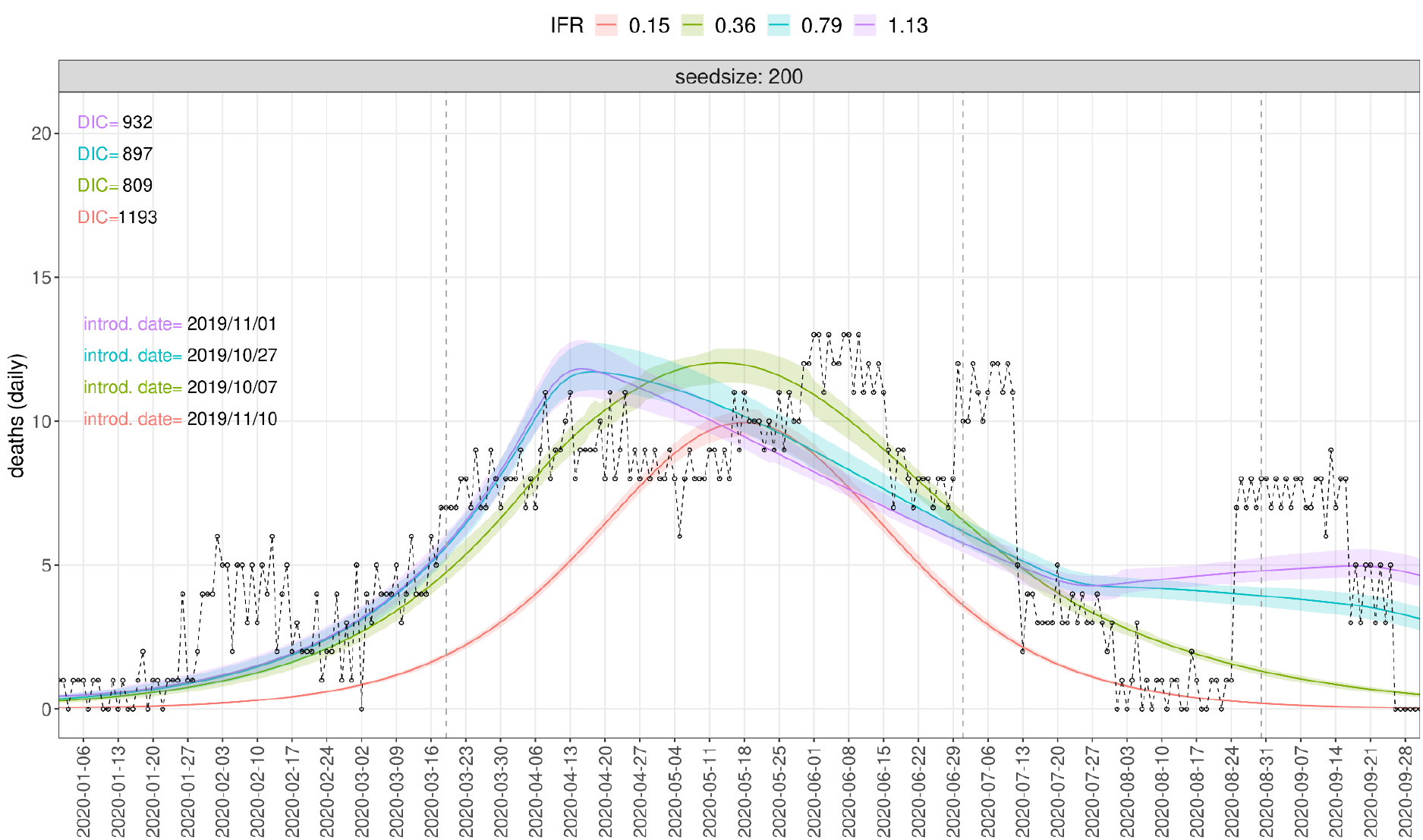
Dynamics generated by sampling the posterior distributions of fitting parameters, at a seed size of 200 and four IFR values from 0.15% to 1.13%. The best fit (lowest DIC value) is at IFR=0.36%. The dashed black line and circles show the daily number of excess burials. Only the period from 23 February to 24 August was used for fitting.

Due to the 3-week delay from infection to death (Figure 5) and the relatively low population-average IFR values in Mogadishu’s young population, the early rise in deaths coupled with the very low estimates of *R*_*0*_ lead to early introduction date estimates of October-November 2019 (Figure 5, Table 1). The date of introduction shifts to late November 2019 only if a large seeding event (n=500) is assumed (SI Figure 9).

**Figure 5.**
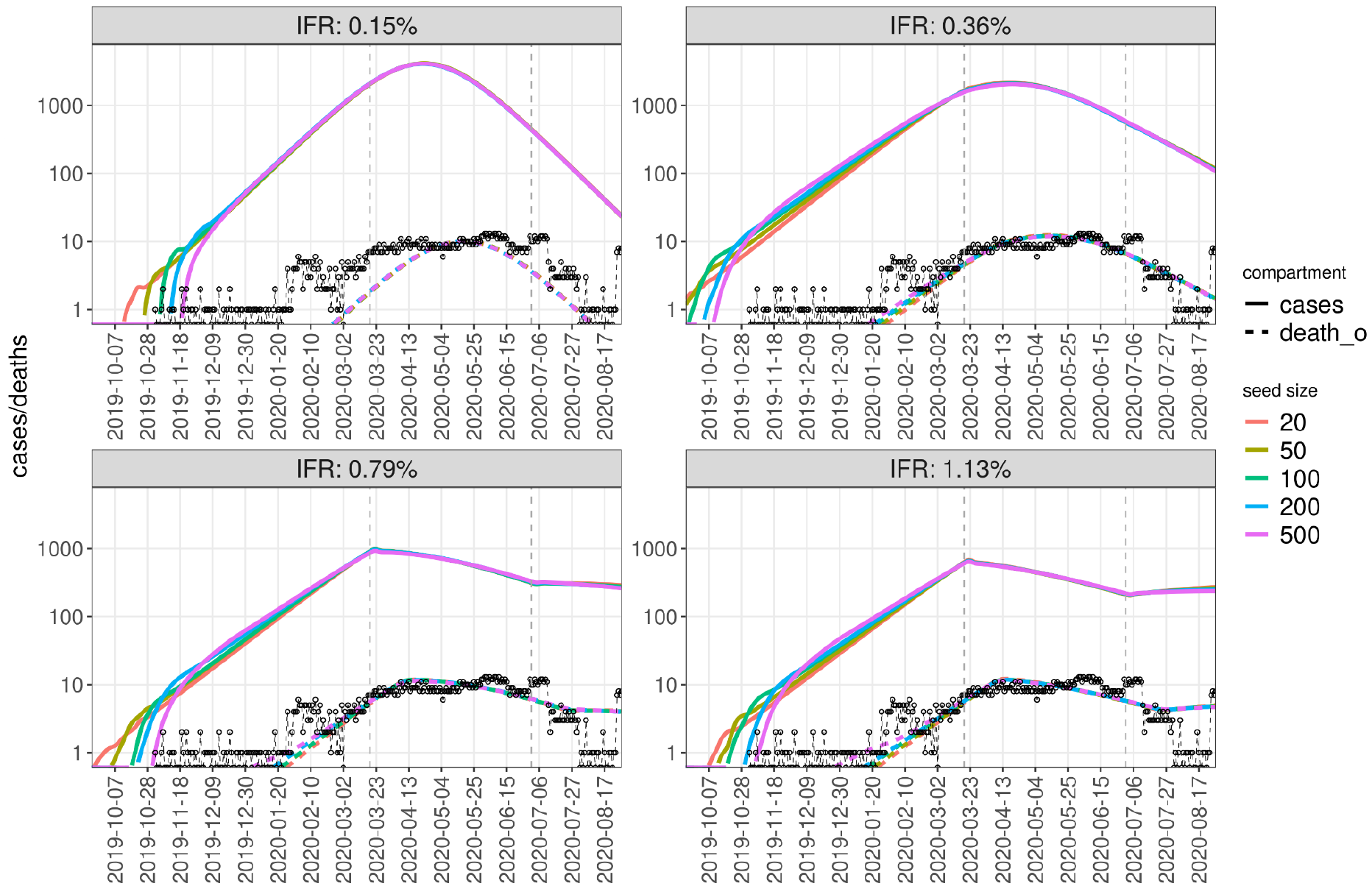
Simulated dynamics of cases (solid lines) and deaths (dashed, colored) for different seed sizes (colors) and IFR values (subplots) using the mean values of fitted parameters, compared to daily number of excess burials (black dashed line and circles).

### Estimates for the effect of NPIs

The estimates for the effectiveness of NPIs show strong positive correlation with the population-average IFR and strong negative correlation with the seed size (Figure 2). For the first NPI period when *StringencyIndex* was the highest (19/March to 30/June) we obtained median estimates of transmissibility reduction below 1% (0.32-0.4%, *NPI_scale*=0.005-0.007) when using the literature-derived IFR estimate. The NPI-caused transmissibility reduction is 10-17% for IFR=0.36%, 22-25% for IFR=0.79% and 25-28% for IFR=1.13% (Figure 3). These values are substantially lower than NPI-induced reductions in contact rates in high income countries (30), however they have an effect on the growth of cases for the fits with IFR values of 0.79% and above, as they break the exponential growth of cases and result in the long plateau of deaths that we observe in our burial data (Figure 5). This breakpoint in the dynamics of infections due to the stronger effect of NPIs that we obtain for the fits with IFR above 0.36% shifts the date of introduction to a later date by approximately three weeks.

### Uncertainty in estimates due to parameter correlations

There are strong positive correlations between all three fitting parameters (SI Figure 11), nevertheless 50% and 95% confidence intervals for all three are relatively narrow (Figure 2). A higher assumed IFR value and a larger seed size both shift the date of introduction to later dates, while lowering the estimates for the basic reproduction number (Figure 2-3). The best fits in terms of DIC values are obtained with intermediate values of the IFR (Figure 3), while larger seed sizes only marginally improve the fit quality for a given IFR while arguably less likely themselves.

### Deaths from political violence in 2020

Since Somalia is heavily affected by political violence, including armed conflicts and terror attacks (31), we also investigated if a rise in violent deaths could explain the sustained rise in burials observed February-July 2020. We analysed the number of fatalities due to political violence documented for Somalia in the Armed Conflict Location & Event Data Project (ACLED) database for the years 2018, 2019 and 2020. Compared to 2018-2019, for the year of 2020 we found no increase, but rather a reduction, in the number of fatalities due to political violence in the Banadir region (SI Figure 4-5). While there was one major terrorist attack claiming 85 lives on the 28th December 2019, this was followed by a long period of deaths below the level of the previous year, followed by two major incidents (resulting in 20 and 26 deaths, respectively) in August 2020. In the period from February to July, when the daily burial rate doubled from its baseline, there was no increase of fatalities due to political violence, therefore it is unlikely the rise in burials could be explained by this exogenous factor.

## Discussion

Fitting excess mortality in Mogadishu from 23/02/2020 by a validated SARS-CoV-2 age-structured compartmental model (22,24) we arrived at date of introduction estimates of October-November 2019, more than two months earlier than previous estimates (32). Additionally, our estimates of the basic reproduction number between 1.3 and 1.5 are also markedly lower than previous estimates (33). These two findings are not only due to the early appearance of excess mortality from late January (and more consistently from late February), but also the slow rise of deaths and their sustained plateau from April to July, leading to the low *R*_*0*_ estimate and a consequent dating-back of the introduction date to very early time points. While the epidemiological model can fit the deaths data relatively well (Figure 4-5), the introduction dates of October-November 2019 and large seed sizes of 100-500 infecteds entering the region are surprising based on the current understanding of the early phase of the COVID-19 pandemic, dating the introduction of the pathogen to Africa around January 2020 (14,34).

This study has several limitations. Our model fitting of excess deaths is predicated on the strong assumption that the unexplained rise in burials from late January 2020 was due to deaths caused by SARS-CoV-2 infection. We investigated a number of alternative hypotheses other than COVID-19 that could explain the observed excess mortality.

There was an ongoing cholera epidemic in Somalia following floods since 2017 (28), resulting in 19 confirmed deaths in Banadir (mostly children) from January to October 2020. In the four weeks of 20 January to 06 February, approximately coinciding with the first transient increase of excess burials in our dataset, there were four confirmed cholera deaths in the whole of Somalia, after a preceding period of no reported deaths. In the period from 16 February to 12 April there were 8 further cholera deaths reported Somalia-wide, and another 12 deaths in June-July. These numbers are much lower than the observed increase of burials between February and October 2020: approximately 1500 excess burials were directly identified from satellite imagery and we estimated total excess deaths in Banadir to be between four and twelve thousand. While some underestimation of cholera-related deaths is possible, due to its well-identifiable pathology we consider it unlikely that a major cholera outbreak was almost entirely missed and could explain a substantial proportion of the excess mortality.

The very early date of introduction estimates are due both to the early rise of excess deaths and the low *R*_*0*_ estimate stemming from the slow rise and long plateau of the curve. It is possible that the real peak of deaths was in fact higher and the curve had a sharper exponential rise, but the number of burials underestimates excess deaths during the peak period of the pandemic, perhaps because of out-migration from the city (as observed in India (35)) during the pandemic or opening of new burial sites not included in our satellite data. While field visits and interviews did not identify new burial sites and we therefore cannot ascertain the veracity of this hypothesis, we nevertheless approximated it by re-fitting the model to the pre-peak period up to 13 April only. This led to a small shift in the date of introduction to a later date, but still resulted in estimates of late October to mid-November for seed sizes of 30 or 100 (SI Figure 12-13), with median *R*_*0*_ estimates rising to 1.4-1.6 for a seed size of 30 (1.3-1.4 for a seed size of 100).

There are two, non-exclusive ways to interpret these findings. On the one hand, given uncertainties about the earliest phase of the pandemic in Wuhan (36), it is possible that SARS-CoV-2 was imported to Mogadishu at a much earlier date than most consensus estimates. Mogadishu, the only international airport in Somalia, received flights with over thirty thousand seats in total per month in the period before the COVID-19 pandemic (34). The country has connecting flights with multiple countries (UAE, Turkey, Kenya, Ethiopia, Qatar) that have several daily flights with China and in two cases (Turkey, UAE) with Wuhan (37). Moreover, news articles reported at least 34 Somali students in Wuhan (38), with the entire Somali diaspora likely to be larger, as trade and general economic relations between China and Somalia have been expanding in the last two decades, resulting in a growing Somali (and other African) diaspora in China (39). Larger neighbouring countries Kenya and Ethiopia have far more extensive trade (40,41) and travel (42,43) flows with China, making indirect importation to Somalia possible, but implying that earlier introduction dates could have happened for those countries as well. SARS-CoV-2 positive routine samples from mid-December 2019 were also found in Italy (16), France (44) and the United States (45), suggesting the pathogen was circulating in small numbers by the end of 2019 outside of China, but not resulting in excess mortality until Spring 2020. Phylogenetic analysis (46) suggests that a progenitor of the SARS-CoV-2 variant first identified in Wuhan might have been spreading outside of China months before the known beginning of the city’s outbreak.

In the compartmental-deterministic framework we used, superspreading events can only be incorporated as static model inputs (ie. an injection of cases into the model), although the large seed sizes used as input parameters can be interpreted as proxies for early superspreading events that followed smaller seeding events. In Somali society, superspreading events such as large funerals or marriages may be more likely than in Europe, such that the importation of even a few seed cases in late 2019 might have resulted in extensive early propagation.

If some of the excess mortality observed in our dataset was due to causes other than COVID-19, a more sharply rising epidemic curve might be hidden within the curve of all excess deaths, which, if disentangled, would lead to a higher *R*_*0*_ estimate and therefore a later date of introduction. Conversely, if a very early introduction did occur, leading to a rise in deaths from February 2020, behavioral adaptation by the general population might have reduced contact rates and resulted in the lower *R*_*0*_. During 2020 Mogadishu was not affected by large-scale armed conflict, influx of displaced people or food insecurity, as in previous phases of the protracted crisis in Somalia. Differential under-ascertainment of burials over time in the satellite imagery analysis may therefore provide a more plausible explanation. Burials in the deceased’s village of origin outside the capital and a potential decrease in the number of these burials (and thereby an increase of burials within city limits) due to mobility restrictions could have also played a role. Either way, it is also plausible that some of the excess mortality is in fact due to the NPIs themselves and other socio-economic disruptions due to the pandemic, though they cannot explain the rise in excess burials starting from February 2020.

Our best fits were obtained at IFR values higher than if age-specific IFRs were identical to consensus estimates from high-income countries. With the above qualifications in mind, this finding can be interpreted as supporting a higher IFR than expected from age demographics only, which could be due to untreated comorbidities and limited access to COVID-19 treatment (47).

The *R*_*0*_ estimates between 1.3-1.5 are substantially lower than consensus estimates for SARS-CoV-2 for China (48,49), Europe (23,50) or the United States (51). We note there is no empirical contact matrix or real-time mobility data available for Somalia, it is therefore possible that contact structures are in reality somewhat different from the projected contact matrix (52) for its neighbour (Ethiopia) that we used for model fitting, contributing to a lower reproduction number. Another possibility is that susceptibility to infection in younger individuals (SI Figure 7) is lower than estimates inferred for middle and high-income countries (22), i.e. that the lower median age in Somalia reduced the *R*_*0*_ further. Relatively high ventilation of houses and proportion of time spent outdoors due to warm weather may also have reduced transmissibility. Other factors such as cross-immunity have also been proposed (53).

Finally, we note that our model fitting resulted in attack rates between 15-50% (Figure 6), which would have left a large pool of susceptibles for a second wave to develop if the reproduction number increased due to introduction of new variants in late 2020. Indeed, a reportedly sharp pandemic wave was observed in Somalia (54) from late February to May 2021.

**Figure 6.**
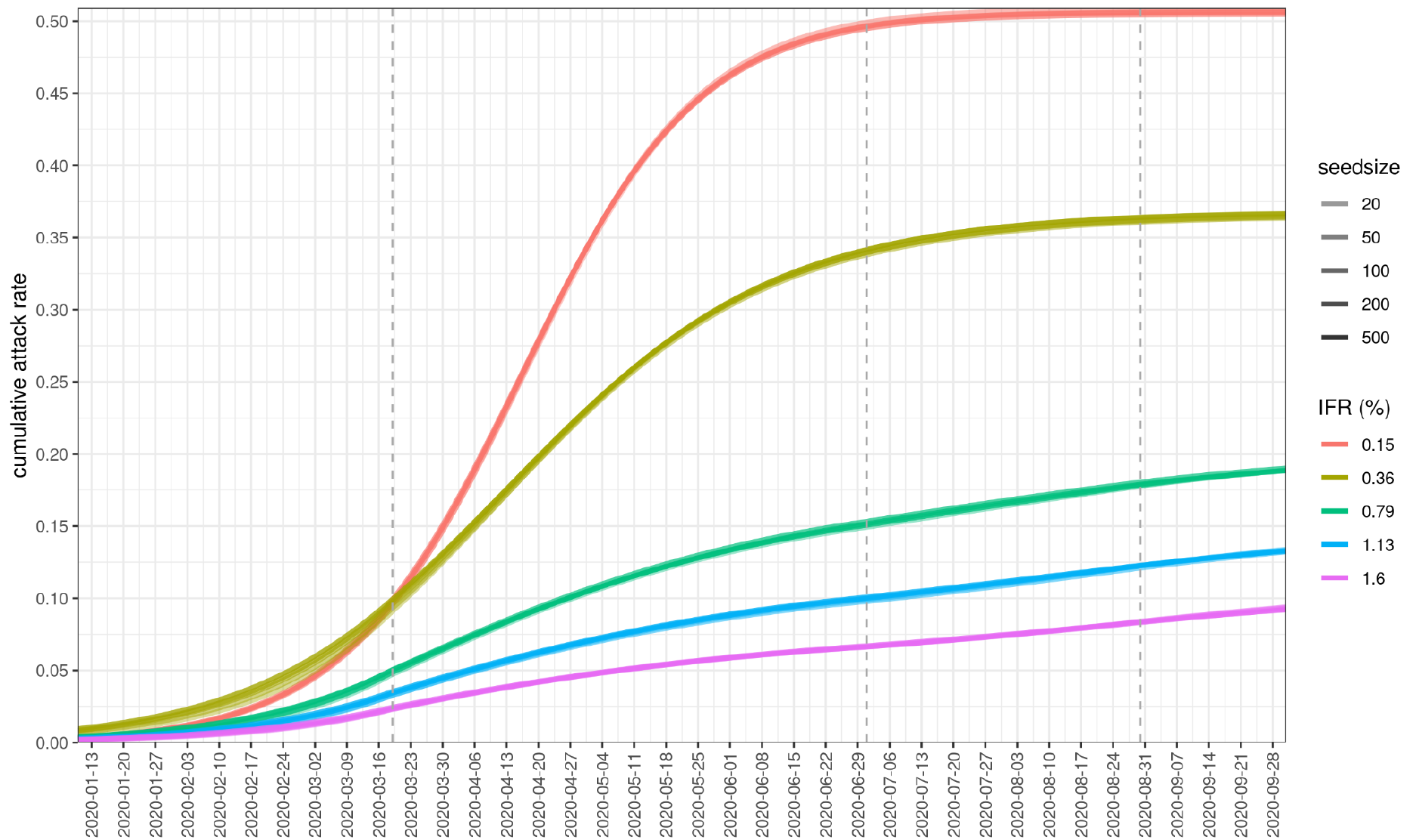
Cumulative attack rates for different seed sizes and IFR values, fitting the period 23/02/2020-24/08/2020. Different IFR values lead to different estimates of *R*_*0*_ and *NPI_scale*, resulting in different herd immunity thresholds and attack rates.

In summary, our analysis was based on the assumption that the rise in excess mortality observed via satellite imagery of cemeteries in Mogadishu early 2020 was due to deaths caused by SARS-CoV-2 infection. Under this assumption, model fitting of the time series of deaths suggests that SARS-CoV-2 could have been introduced to Somalia’s capital substantially earlier than previously thought and had a reproduction number lower than consensus estimates from middle and high income countries, leading to an effective reproduction number around 1 from April to July and a long plateau of excess deaths. If these excess deaths were indeed due to SARS-CoV-2 infections, this raises several questions about the pathogen’s introduction to Africa and the true burden of the pandemic on the continent. Further investigation of mortality trends and SARS-CoV-2 epidemiology in Somalia and other low-income countries is warranted to paint a more conclusive picture, and help to better predict future waves of the pandemic in these settings.

## Data

The data on burials, stringency index, deaths due to political violence and epidemiological parameters used for the CovidM model are available in the github repository: https://github.com/mbkoltai/covid_lmic_model along with the scripts to reproduce figures. SI Figure 1 was produced with the scripts available at https://github.com/francescochecchi/mogadishu_burial_analysis where satellite imagery data are also uploaded.

## Authorship

The following authors were part of the Centre for Mathematical Modelling of Infectious Disease COVID-19 Working Group: Adam J Kucharski, Sebastian Funk, Rosalind M Eggo, Mark Jit, W John Edmunds, Amy Gimma, Stefan Flasche, Billy J Quilty, Samuel Clifford, Sam Abbott, James D Munday, Nikos I Bosse, Joel Hellewell, Hamish P Gibbs, Yang Liu, Nicholas G. Davies, Timothy W Russell, Christopher I Jarvis, Alicia Rosello, Carl A B Pearson, Kiesha Prem, Graham Medley, Gwenan M Knight, Akira Endo, Simon R Procter, C Julian Villabona-Arenas, Damien C Tully, Katherine E. Atkins, Sophie R Meakin, Rachel Lowe, Matthew Quaife, Oliver Brady, Kaja Abbas, Rosanna C Barnard, Frank G Sandmann, Kathleen O’Reilly, Mihaly Koltai, Yalda Jafari, William Waites, David Hodgson, Emilie Finch, Ciara V McCarthy, Rachael Pung, Paul Mee, Lloyd A C Chapman, Fiona Yueqian Sun, Stéphane Hué, Kerry LM Wong.

Each contributed in processing, cleaning and interpretation of data, interpreted findings, contributed to the manuscript, and approved the work for publication.

## Supporting information

The Supporting Information contains SI Table 1 and SI Figures 1-15.

## References

1. Johns Hopkins Coronavirus Resource Center [Internet]. Johns Hopkins Coronavirus Resource Center. [cited 2021 Mar 31]. Available from: https://coronavirus.jhu.edu/region

2. Levin AT, Meyerowitz-Katz G, Owusu-Boaitey N, Cochran KB, Walsh SP. Assessing the Age Specificity of Infection Fatality Rates for COVID-19: Systematic Review, Meta-Analysis, and Public Policy Implications. :29.

3. Favas C, Jarrett P, Ratnayake R, Watson OJ, Checchi F. Country differences in transmissibility, age distribution and case-fatality of SARS-CoV-2: a global ecological analysis. medRxiv. 2021 Feb 19;2021.02.17.21251839.

4. Report 34 - COVID-19 Infection Fatality Ratio Estimates from Seroprevalence [Internet]. Imperial College London. [cited 2021 Mar 31]. Available from: http://www.imperial.ac.uk/medicine/departments/school-public-health/infectious-disease-epidemiology/mrc-global-infectious-disease-analysis/covid-19/report-34-ifr/

5. Biccard BM, Joubert I, Kifle F, Kluyts H-L, Macleod K, Mekonnen Z, et al. Patient care and clinical outcomes for patients with COVID-19 infection admitted to African high-care or intensive care units (ACCCOS): a multicentre, prospective, observational cohort study. The Lancet. 2021 May 22;397(10288):1885–94.

6. van Zandvoort K, Jarvis CI, Pearson CAB, Davies NG, Covid C. Response strategies for COVID-19 epidemics in African settings: a mathematical modelling study. 2020;39.

7. Covid: Does Tanzania have a hidden epidemic? BBC News [Internet]. 2021 Mar 17 [cited 2021 Mar 31]; Available from: https://www.bbc.com/news/56242358

8. Wiens KE, Mawien PN, Rumunu J, Slater D, Jones FK, Moheed S, et al. Seroprevalence of anti-SARS-CoV-2 IgG antibodies in Juba, South Sudan: a population-based study. medRxiv. 2021 Mar 12;2021.03.08.21253009.

9. Gomaa MR, Rifay ASE, Shehata M, Kandeil A, Kamel MN, Marouf MA, et al. Incidence, household transmission, and neutralizing antibody seroprevalence of Coronavirus Disease 2019 in Egypt: Results of a community-based cohort. PLOS Pathog. 2021 Mar 11;17(3):e1009413.

10. Mwananyanda L, Gill CJ, MacLeod W, Kwenda G, Pieciak R, Mupila Z, et al. COVID-19 deaths detected in a systematic post-mortem surveillance study in Africa. medRxiv. 2020 Dec 24;2020.12.22.20248327.

11. Van Elsland S, Watson O, Alhaffar M, Mehchy Z, Whittaker C, Akil Z, et al. Report 31: Estimating the burden of COVID-19 in Damascus, Syria: an analysis of novel data sources to infer mortality under-ascertainment [Internet]. Imperial College London; 2020 Sep [cited 2021 Mar 31]. Available from: http://spiral.imperial.ac.uk/handle/10044/1/82443

12. Karlinsky A, Kobak D. The World Mortality Dataset: Tracking excess mortality across countries during the COVID-19 pandemic. medRxiv. 2021 Jan 29;2021.01.27.21250604.

13. Watson OJ, Alhaffar M, Mehchy Z, Whittaker C, Akil Z, Brazeau NF, et al. Leveraging community mortality indicators to infer COVID-19 mortality and transmission dynamics in Damascus, Syria. Nat Commun. 2021 Apr 22;12(1):2394.

14. Adepoju P. Nigeria responds to COVID-19; first case detected in sub-Saharan Africa. Nat Med. 2020 Mar 11;26(4):444–8.

15. Nadeau SA, Vaughan TG, Scire J, Huisman JS, Stadler T. The origin and early spread of SARS-CoV-2 in Europe. Proc Natl Acad Sci [Internet]. 2021 Mar 2 [cited 2021 Mar 31];118(9). Available from: https://www.pnas.org/content/118/9/e2012008118

16. La Rosa G, Mancini P, Bonanno Ferraro G, Veneri C, Iaconelli M, Bonadonna L, et al. SARS-CoV-2 has been circulating in northern Italy since December 2019: Evidence from environmental monitoring. Sci Total Environ. 2021 Jan 1;750:141711.

17. Koum Besson E, Norris A, Bin Ghouth AS, Freemantle T, Alhaffar M, Vazquez Y, et al. Excess mortality during the COVID-19 pandemic in Aden governorate, Yemen: a geospatial and statistical analysis [Internet]. Epidemiology; 2020 Oct [cited 2021 Apr 28]. Available from: http://medrxiv.org/lookup/doi/10.1101/2020.10.27.20216366

18. Warsame A, Bashiir F, Freemantle T, Williams C, Vazquez Y, Reeve C, et al. Excess mortality during the COVID-19 pandemic: a geospatial and statistical analysis in Mogadishu, Somalia. medRxiv. 2021 May 17;2021.05.15.21256976.

19. Division UNP. wpp2019: World Population Prospects 2019 [Internet]. 2020.[cited 2021 Apr 27]. Available from: https://CRAN.R-project.org/package=wpp2019

20. Qader SH, Lefebvre V, Tatem AJ, Pape U, Jochem W, Himelein K, et al. Using gridded population and quadtree sampling units to support survey sample design in low-income settings. Int J Health Geogr. 2020 Mar 26;19(1):10.

21. Warsame A, Frison S, Gimma A, Checchi F. Retrospective estimation of mortality in Somalia, 2014-2018: a statistical analysis. :35.

22. Davies NG, Klepac P, Liu Y, Prem K, Jit M, Eggo RM. Age-dependent effects in the transmission and control of COVID-19 epidemics. Nat Med. 2020 Aug;26(8):1205–11.

23. Davies NG, Kucharski AJ, Eggo RM, Gimma A, Edmunds WJ, Jombart T, et al. Effects of non-pharmaceutical interventions on COVID-19 cases, deaths, and demand for hospital services in the UK: a modelling study. Lancet Public Health. 2020 Jul 1;5(7):e375–85.

24. Sandmann FG, Davies NG, Vassall A, Edmunds WJ, Jit M, Sun FY, et al. The potential health and economic value of SARS-CoV-2 vaccination alongside physical distancing in the UK: a transmission model-based future scenario analysis and economic evaluation. Lancet Infect Dis [Internet]. 2021 Mar 18 [cited 2021 Apr 27]; Available from: https://www.sciencedirect.com/science/article/pii/S1473309921000797

25. Cao B, Wang Y, Wen D, Liu W, Wang J, Fan G, et al. A Trial of Lopinavir–Ritonavir in Adults Hospitalized with Severe Covid-19. N Engl J Med. 2020 May 7;382(19):1787–99.

26. Linton NM, Kobayashi T, Yang Y, Hayashi K, Akhmetzhanov AR, Jung S, et al. Incubation Period and Other Epidemiological Characteristics of 2019 Novel Coronavirus Infections with Right Truncation: A Statistical Analysis of Publicly Available Case Data. J Clin Med. 2020 Feb;9(2):538.

27. COVID-19 Government Response Tracker [Internet]. [cited 2021 Mar 30]. Available from: https://www.bsg.ox.ac.uk/research/research-projects/covid-19-government-response-tracker

28. WHO EMRO | Outbreak update – Cholera in Somalia, 27 December 2020 | Cholera | Epidemic and pandemic diseases [Internet]. [cited 2021 Apr 28]. Available from: http://www.emro.who.int/pandemic-epidemic-diseases/cholera/outbreak-update-cholera-in-somalia-27-december-2020.html

29. Sanche S, Lin YT, Xu C, Romero-Severson E, Hengartner N, Ke R. High Contagiousness and Rapid Spread of Severe Acute Respiratory Syndrome Coronavirus 2 - Volume 26, Number 7—July 2020 - Emerging Infectious Diseases journal - CDC. [cited 2021 Apr 27]; Available from: https://www.nc.cdc.gov/eid/article/26/7/20-0282_article

30. Jarvis CI, Van Zandvoort K, Gimma A, Prem K, CMMID COVID-19 working group, Klepac P, et al. Quantifying the impact of physical distance measures on the transmission of COVID-19 in the UK [Internet]. Epidemiology; 2020 Apr [cited 2021 Feb 18]. Available from: http://medrxiv.org/lookup/doi/10.1101/2020.03.31.20049023

31. ACLED 2020: The Year in Review [Internet]. 2021 [cited 2021 Apr 28]. Available from: https://acleddata.com/2021/03/18/acled-2020-the-year-in-review/

32. Menkir TF, Chin T, Hay JA, Surface ED, De Salazar PM, Buckee CO, et al. Estimating internationally imported cases during the early COVID-19 pandemic. Nat Commun. 2021 Jan 12;12(1):311.

33. Musa SS, Zhao S, Wang MH, Habib AG, Mustapha UT, He D. Estimation of exponential growth rate and basic reproduction number of the coronavirus disease 2019 (COVID-19) in Africa. Infect Dis Poverty. 2020 Jul 16;9(1):96.

34. Rice BL, Annapragada A, Baker RE, Bruijning M, Dotse-Gborgbortsi W, Mensah K, et al. Variation in SARS-CoV-2 outbreaks across sub-Saharan Africa. Nat Med. 2021 Mar;27(3):447–53.

35. Sapra I, Nayak BP. The protracted exodus of migrants from Hyderabad in the time of COVID-19. J Soc Econ Dev [Internet]. 2021 May 6 [cited 2021 Jun 14]; Available from: https://doi.org/10.1007/s40847-021-00155-z

36. Bloom JD, Chan YA, Baric RS, Bjorkman PJ, Cobey S, Deverman BE, et al. Investigate the origins of COVID-19. Science. 2021 May 14;372(6543):694–694.

37. National Contingency Plan for Preparedness and Response to Coronavirus (COVID-19)-Somalia | HumanitarianResponse [Internet]. [cited 2021 Apr 28]. Available from: https://www.humanitarianresponse.info/en/operations/somalia/document/national-contingency-plan-preparedness-and-response-coronavirus-covid-19

38. Somali Students in Wuhan Speak of Fear, Loneliness, Hunger | Voice of America - English [Internet]. [cited 2021 Apr 28]. Available from: https://www.voanews.com/science-health/coronavirus-outbreak/somali-students-wuhan-speak-fear-loneliness-hunger

39. Lochery E. Somali Ventures in China: Trade and Mobility in a Transnational Economy. Afr Stud Rev. 2020 Mar;63(1):93–116.

40. China (CHN) and Kenya (KEN) Trade [Internet]. [cited 2021 May 4]. Available from: https://oec.world/en/profile/bilateral-country/chn/partner/ken

41. Venkataraman M, Gofie SM. The dynamics of China-Ethiopia trade relations: economic capacity, balance of trade & trade regimes. Bdg J Glob South. 2015 Feb 5;2(1):1–17.

42. Kenya Airways to resume Guangzhou; we examine this market and connections over Nairobi [Internet]. anna.aero. 2020 [cited 2021 May 4]. Available from: https://www.anna.aero/2020/08/11/kenya-airways-to-resume-guangzhou-we-examine-this-market-and-connections-over-nairobi/

43. CAPA Live: Ethiopian Airlines cash positive. 50 flights/week to China | CAPA [Internet]. [cited 2021 May 4]. Available from: https://centreforaviation.com/analysis/reports/capa-live-ethiopian-airlines-cash-positive-50-flightsweek-to-china-551683

44. Deslandes A, Berti V, Tandjaoui-Lambotte Y, Alloui C, Carbonnelle E, Zahar JR, et al. SARS-CoV-2 was already spreading in France in late December 2019. Int J Antimicrob Agents. 2020 Jun 1;55(6):106006.

45. Basavaraju SV, Patton ME, Grimm K, Rasheed MAU, Lester S, Mills L, et al. Serologic Testing of US Blood Donations to Identify Severe Acute Respiratory Syndrome Coronavirus 2 (SARS-CoV-2)–Reactive Antibodies: December 2019–January 2020. Clin Infect Dis [Internet]. 2020 Nov 30 [cited 2021 May 25];(ciaa1785). Available from: https://doi.org/10.1093/cid/ciaa1785

46. Kumar S, Tao Q, Weaver S, Sanderford M, Caraballo-Ortiz MA, Sharma S, et al. An evolutionary portrait of the progenitor SARS-CoV-2 and its dominant offshoots in COVID-19 pandemic. Mol Biol Evol [Internet]. 2021 May 4 [cited 2021 May 25];(msab118). Available from: https://doi.org/10.1093/molbev/msab118

47. Somalia: Decline in primary health care visits and childhood vaccinations during COVID-19 [Internet]. International Committee of the Red Cross. 2020 [cited 2021 Apr 28]. Available from: https://www.icrc.org/en/document/somalia-sharp-decline-primary-health-care-visits-and-childhood-vaccinations-during-covid-19

48. Riou J, Althaus CL. Pattern of early human-to-human transmission of Wuhan 2019 novel coronavirus (2019-nCoV), December 2019 to January 2020. Eurosurveillance. 2020 Jan 30;25(4):2000058.

49. Kucharski AJ, Russell TW, Diamond C, Liu Y, Edmunds J, Funk S, et al. Early dynamics of transmission and control of COVID-19: a mathematical modelling study. Lancet Infect Dis. 2020 May 1;20(5):553–8.

50. Flaxman S, Mishra S, Gandy A, Unwin HJT, Mellan TA, Coupland H, et al. Estimating the effects of non-pharmaceutical interventions on COVID-19 in Europe. Nature. 2020 Aug;584(7820):257–61.

51. Ke R, Romero-Severson E, Sanche S, Hengartner N. Estimating the reproductive number R0 of SARS-CoV-2 in the United States and eight European countries and implications for vaccination. J Theor Biol. 2021 May 21;517:110621.

52. Prem K, Zandvoort K van, Klepac P, Eggo RM, Davies NG, Group C for the MM of IDC-19 W, et al. Projecting contact matrices in 177 geographical regions: an update and comparison with empirical data for the COVID-19 era. medRxiv. 2020 Jul 28;2020.07.22.20159772.

53. Organization (WMO) WM, World Meteorological Organization (WMO). First Report of the WMO COVID-19 Task Team : Review on Meteorological and Air Quality Factors Affecting the COVID-19 Pandemic (WMO-No. 1262). Geneva: WMO; 2021. 42 p. (WMO).

54. Salyer SJ, Maeda J, Sembuche S, Kebede Y, Tshangela A, Moussif M, et al. The first and second waves of the COVID-19 pandemic in Africa: a cross-sectional study. The Lancet. 2021 Apr 3;397(10281):1265–75.

