## Supplemental Information for "Date of introduction and epidemiologic patterns of SARS-CoV-2 in Mogadishu, Somalia: estimates from transmission modelling of 2020 excess mortality data"

**Table 1:** parameter values used in the model.

| Parameter | Description | Value |
|---|---|---|
| $N$ | Total population size | 2.2e6 |
| $\Delta t$ | Time step for simulations | 0.25 day |
| $d_E$ | Latent period in days | ~ gamma(μ=3, k=4) |
| $d_P$ | Duration of pre-symptomatic infectiousness in days | ~ gamma(μ=2.1, k=4) |
| $d_C$ | Duration of symptomatic infectiousness in days | ~ gamma(μ=2.9, k=4) |
| $d_S$ | Duration of asymptomatic infectiousness in days | ~ gamma(μ=5, k=4) |
| $y_i$ | Clinical fraction: probability of becoming a symptomatic case if infected for age group *i* | Age-dependent, from (22), SI Figure 7 |
| $u_i$ | Susceptibility to infection. | Age-dependent, from (22), SI Figure 7 |
| $R_0$ | Basic reproduction number (prior value) | 3 |

**Supplementary Figures**

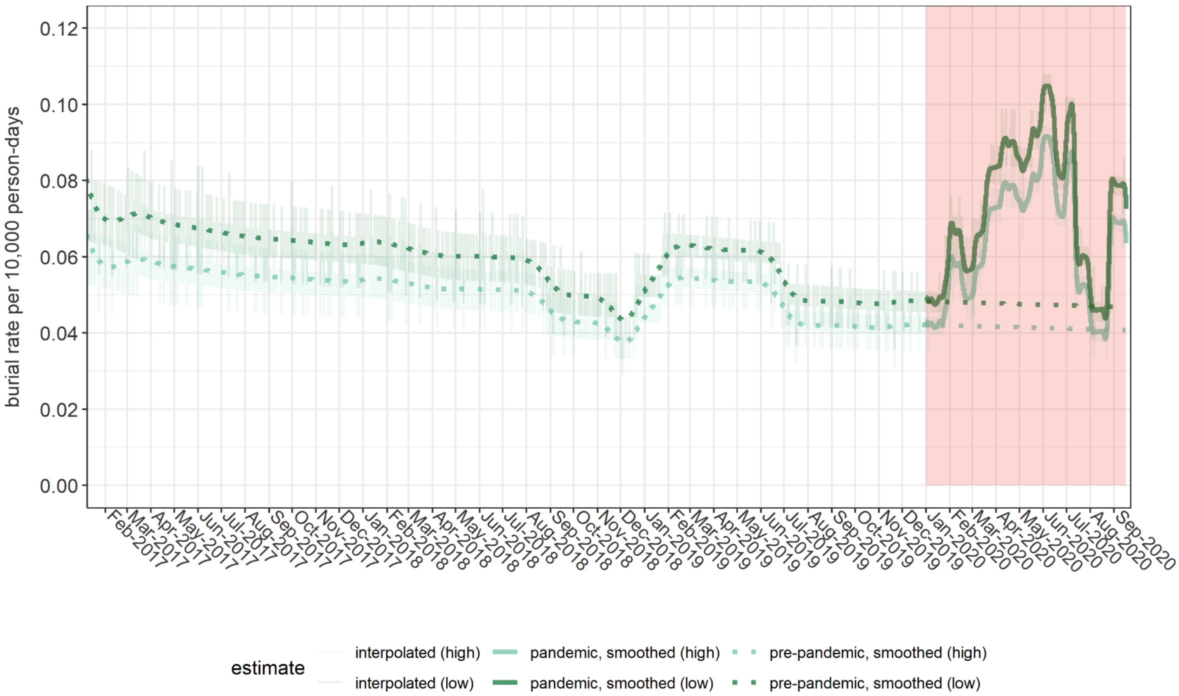


**SI Figure 1**. Burial rate per 10.000 person-days inferred from satellite imagery by interpolating between data points provided by satellite images (for details of methods see the accompanying paper (18)). The color shading represents estimates with low and high population denominators.

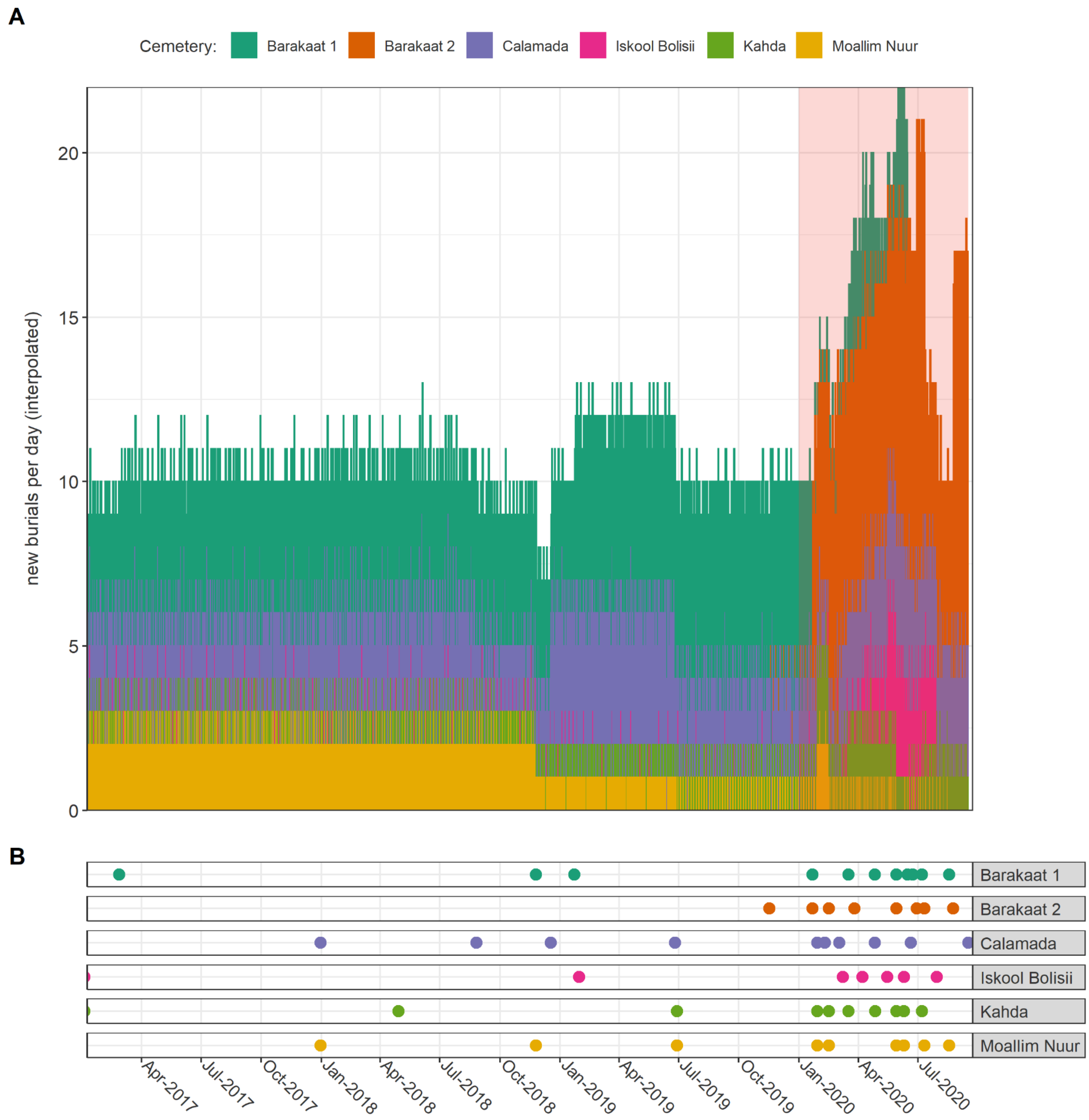


**SI Figure 2**. **A.** Burials by cemetery. Barakaat 1 cemetery was mostly filled up by January 2020 and replaced by its extension Barakaat 2. **B.** Dates for which satellite imagery was acquired.

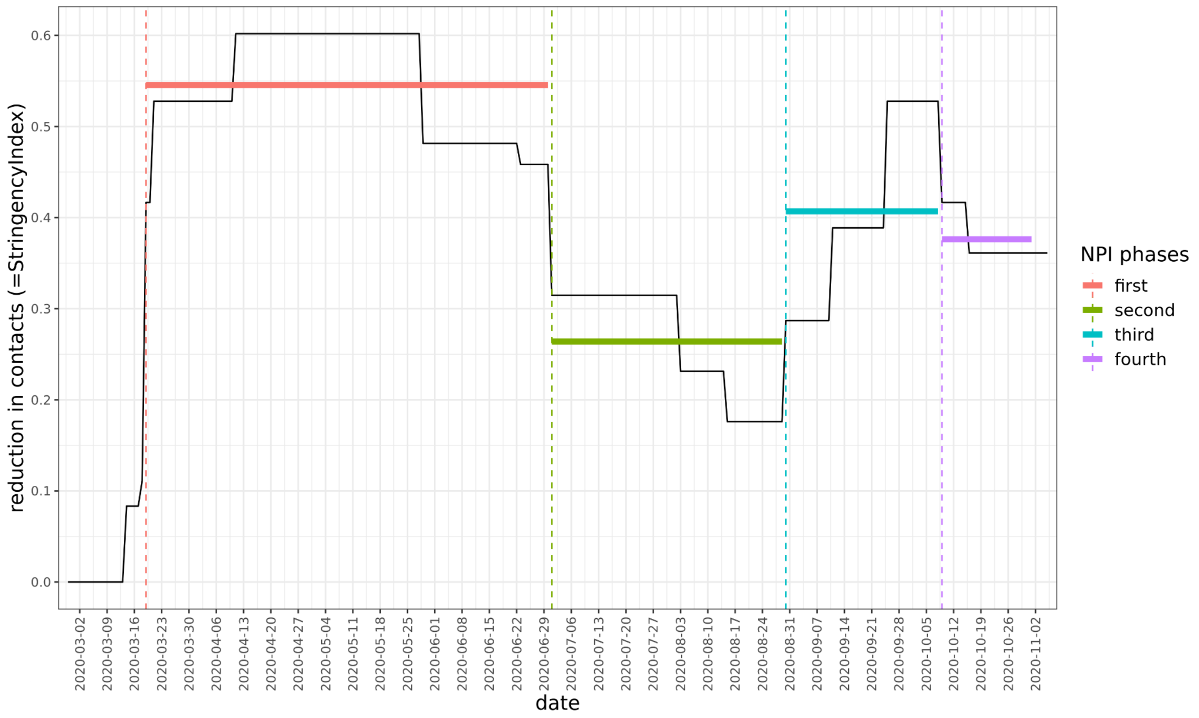


**SI Figure 3**. '*StringencyIndex*' from the Oxford COVID-19 Government Response Tracker database. We took the average (solid horizontal line) of each of the four periods separated by the dashed vertical lines. The fourth period is outside the time window of model fitting. The values are equal to the relative reduction in contacts (transmissibility) if the scaling parameter (*NPI_scale*) is 1.

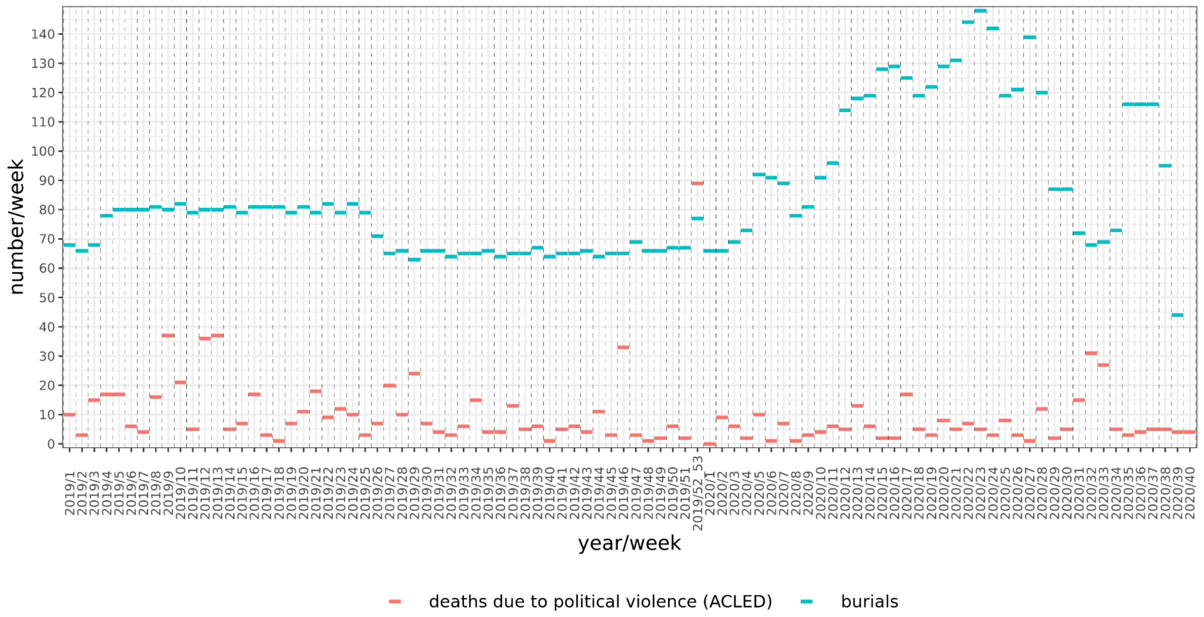


**SI Figure 4**. Weekly fatalities due to political violence in the Banadir region (from ACLED database) compared to the number of burials, going back to January 2019. The infrequency of satellite imagery analysed before 2020 does not enable identification of acute peaks in burials, as would follow a mass casualty incident.

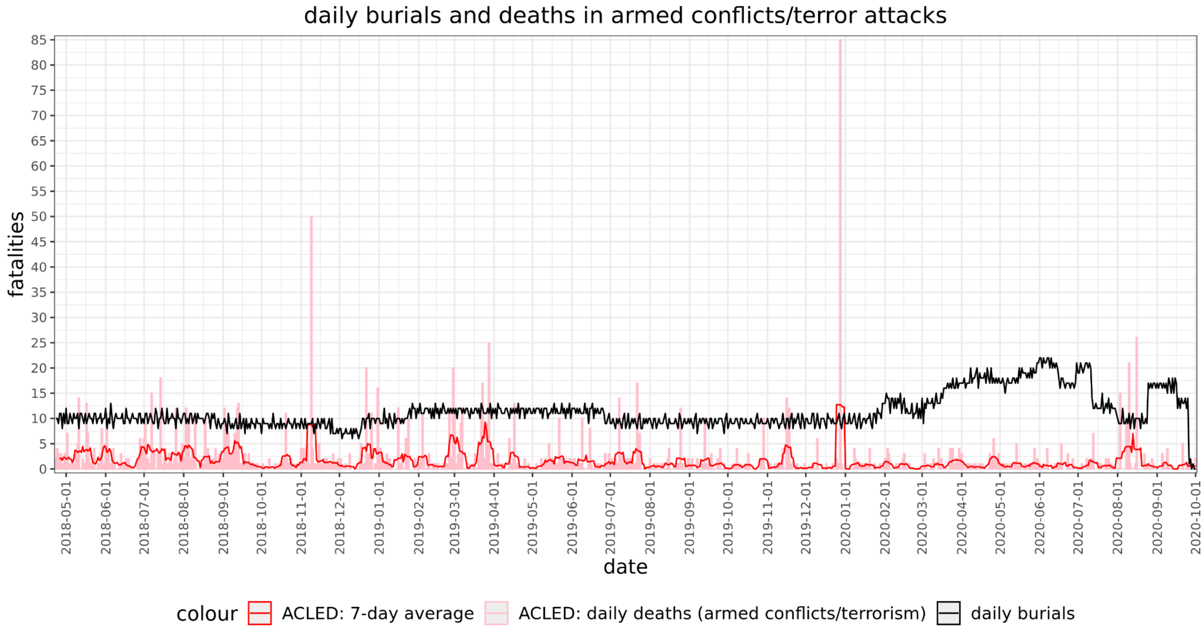


**SI Figure 5**. Daily number of burials compared to the number of deaths due to political violence, between May 2018 and October 2020.

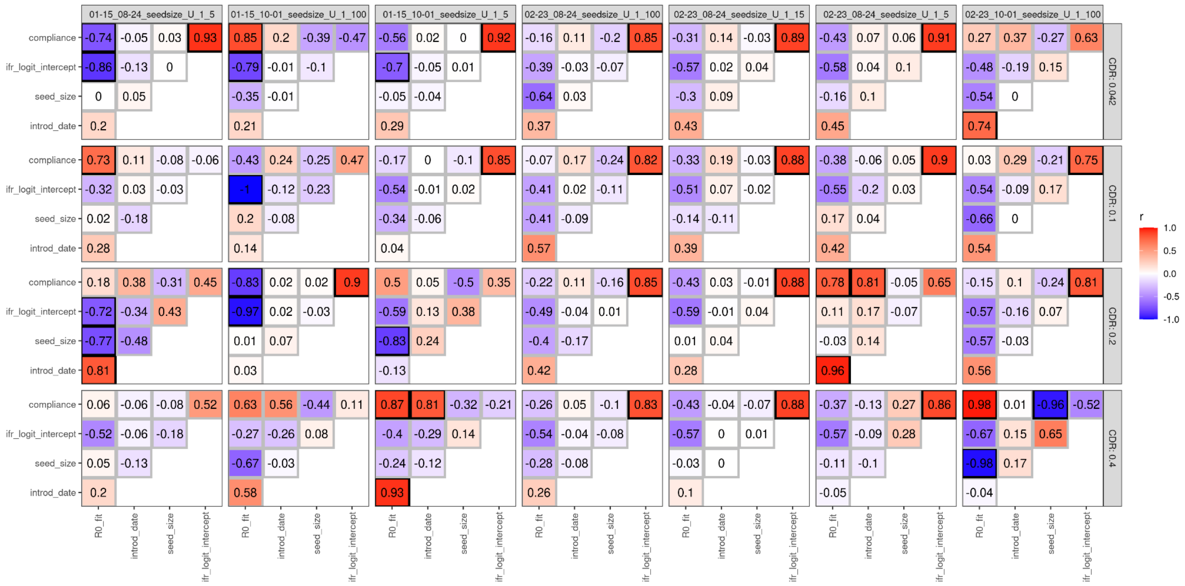


**SI Figure 6**. Correlations between the posterior estimates of all five fitting parameters. Columns are separate fits with different time windows and priors for seed size.

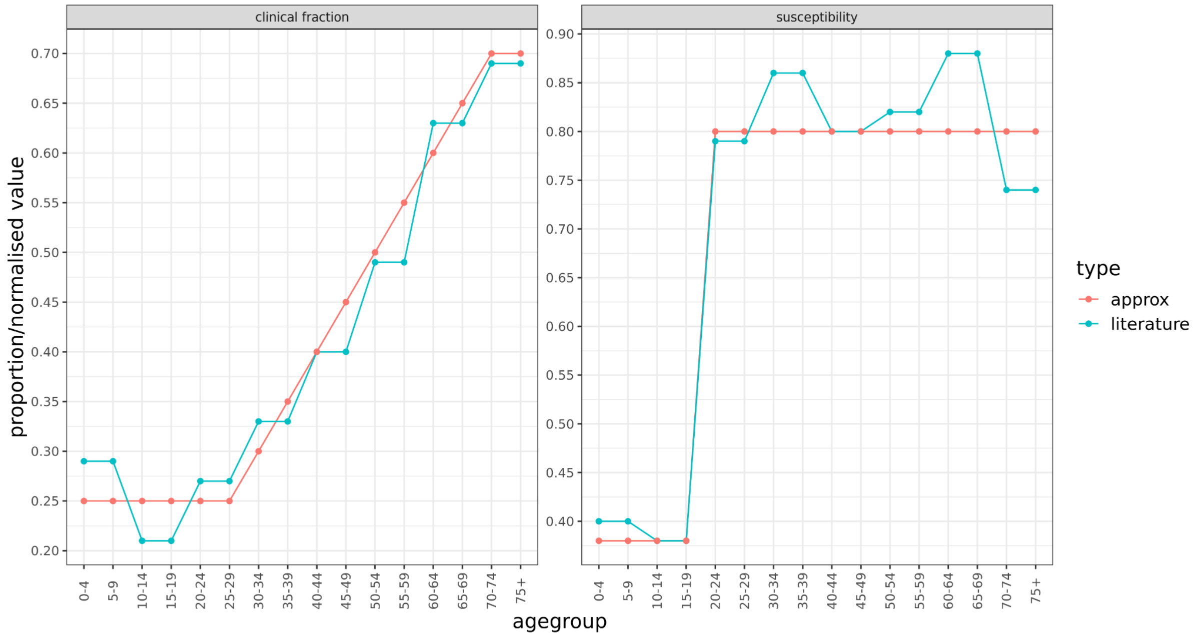


**SI Figure 7**. Clinical fraction and susceptibility by age group. Values from the literature (22) (green) were approximated by a piecewise linear function (red) with a minimum and maximum value and a line connecting them. The linearly approximated values were used as model parameters.

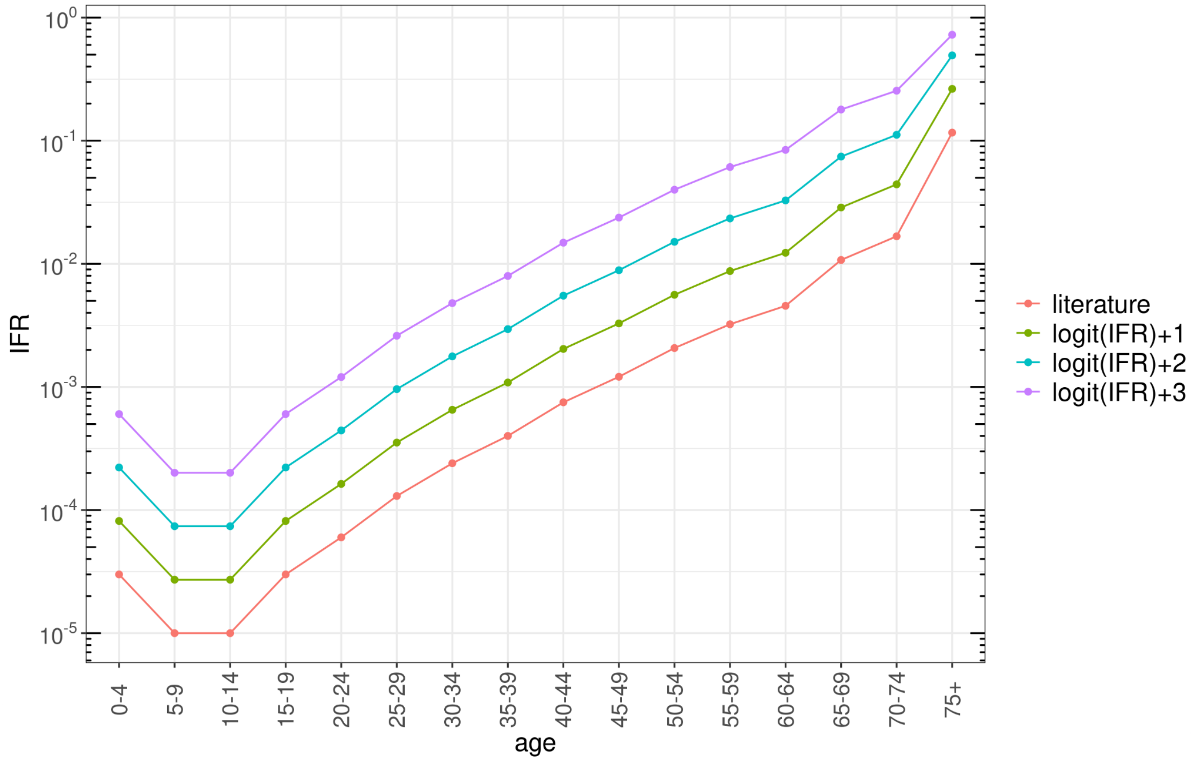


**SI Figure 8**. IFR estimates by age groups, using estimates from (24). Adjusted curves were calculated by taking the logit of the age-specific IFRs and adding the values 1, 2, 3.

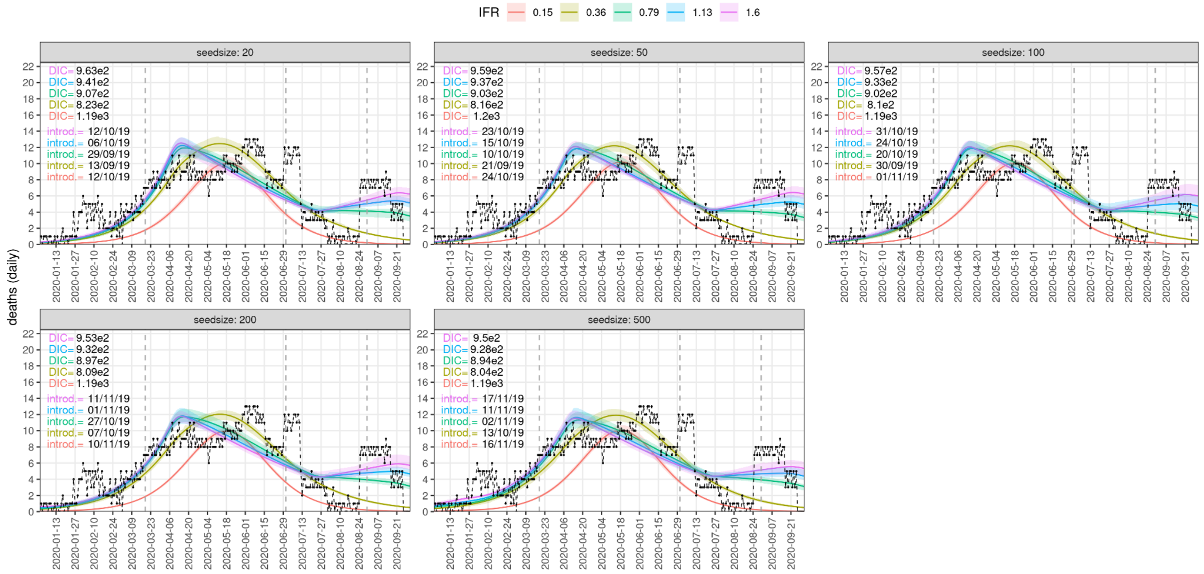


**SI Figure 9**. Dynamic fits with all seed sizes and IFR estimates, fitting the period 23 February to 24 August 2020. Labels show DIC values and median estimates for the date of introduction by population-average IFR values.

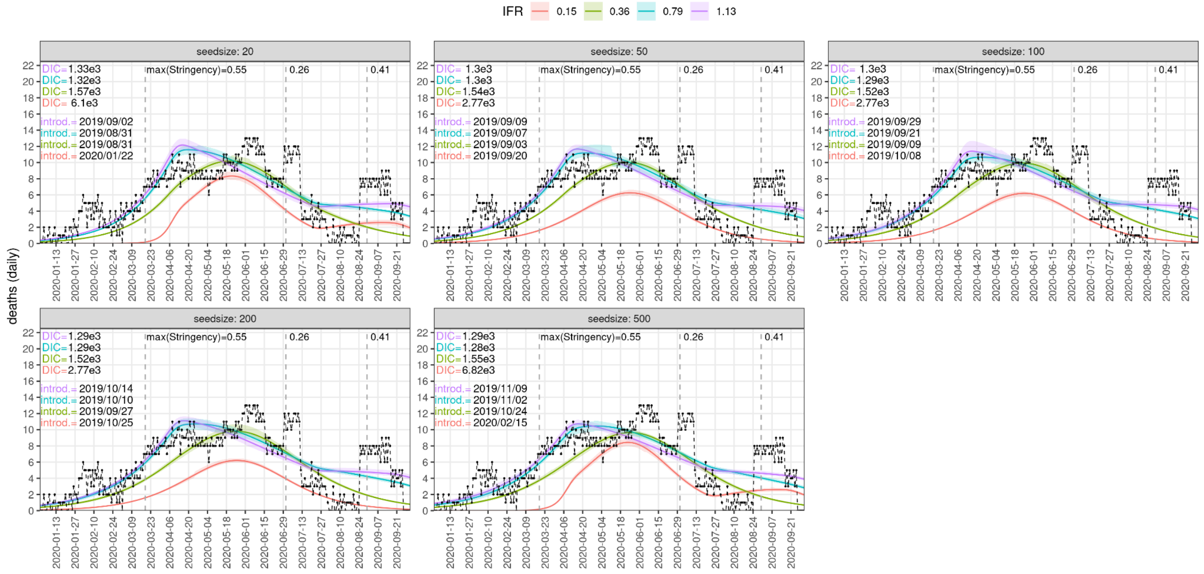


**SI Figure 10**. Dynamic fits with all seed sizes and IFR estimates, fitting the period 15/January-01/October 2020. Labels show DIC values and median estimates for the date of introduction by population-average IFR values. Importations were 20-30 year olds for these simulations.

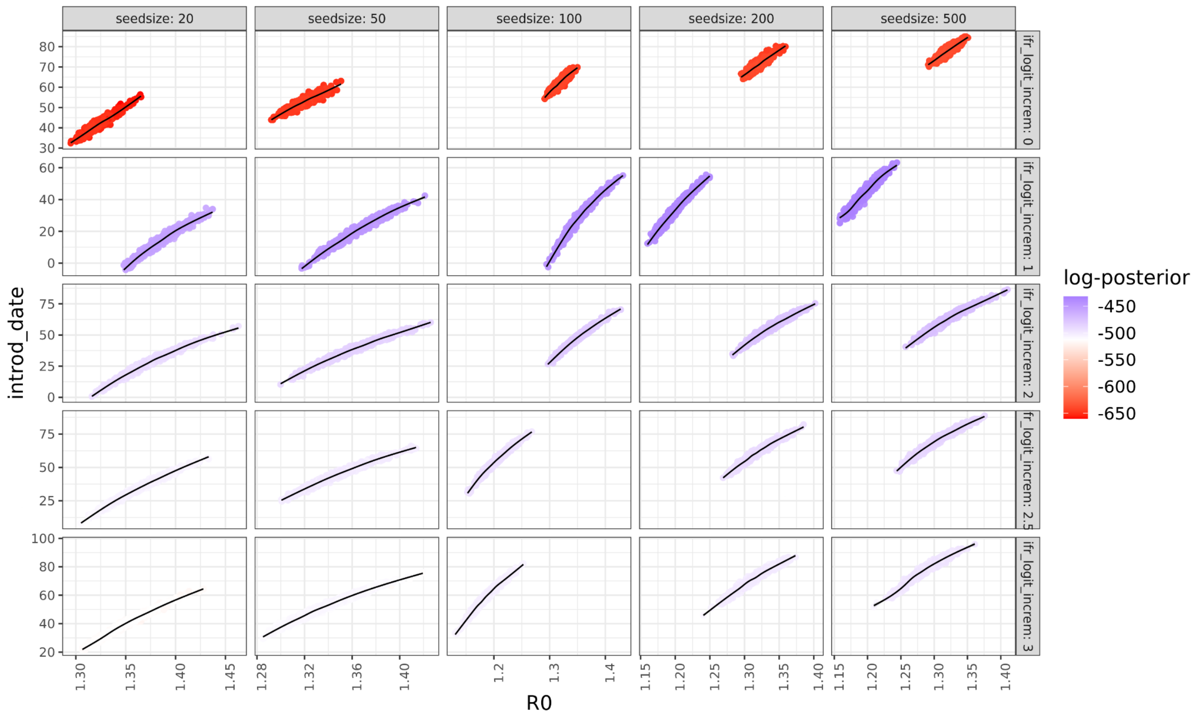


**SI Figure 11**. $R_0$ and introduction date values from their posterior distributions generated by MCMC fitting, showing correlations between the two parameters.

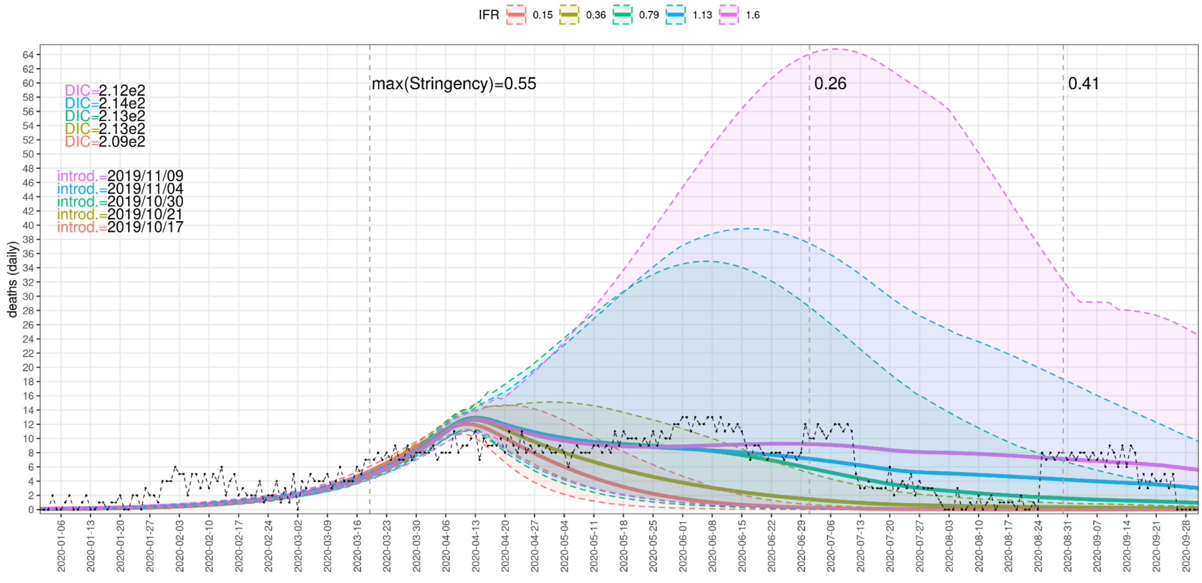


**SI Figure 12**. Model fitting restricted to the period 23/February-13/April, with a seed size of 30. Importations were 20-30 year olds for these fits.

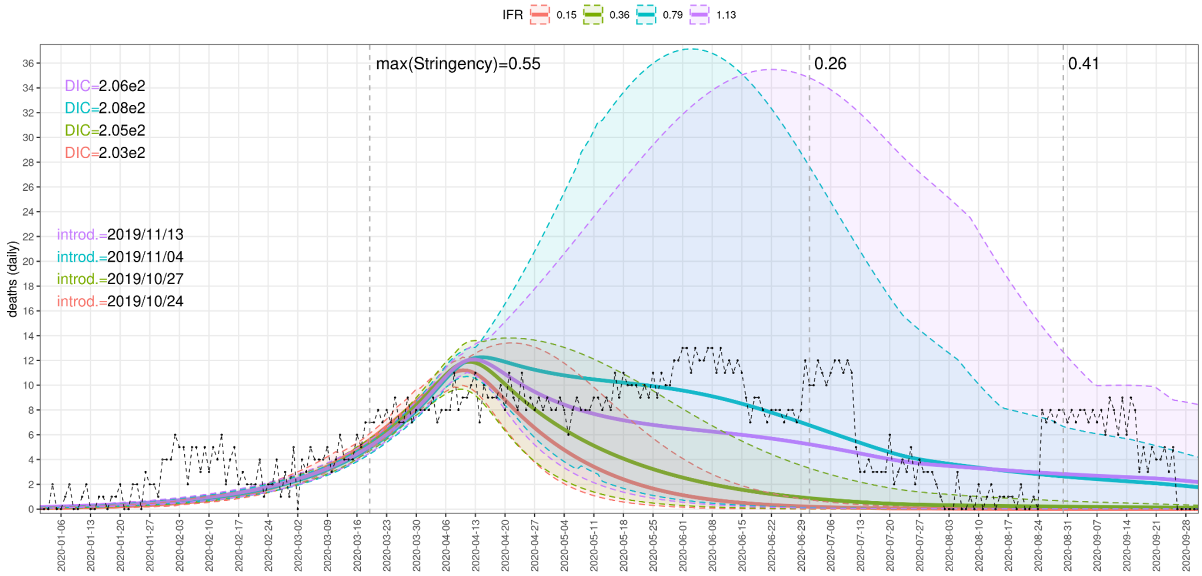


**SI Figure 13**. Model fitting restricted to the period 23/February-13/April, with a seed size of 100.

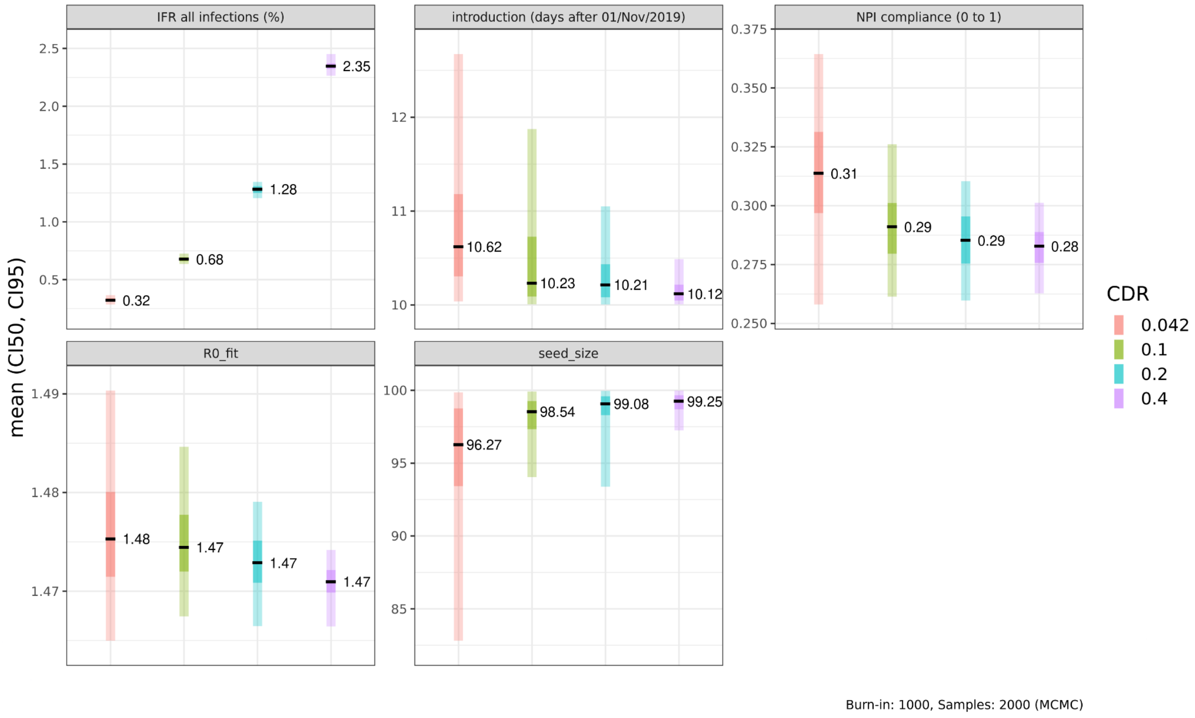


**SI Figure 14**. Posterior distributions for five fitting parameters at different levels of scaling the number of simulated deaths (to different CDR estimates). For these fits the earliest allowed date for the date of introduction was 11/November/2019.

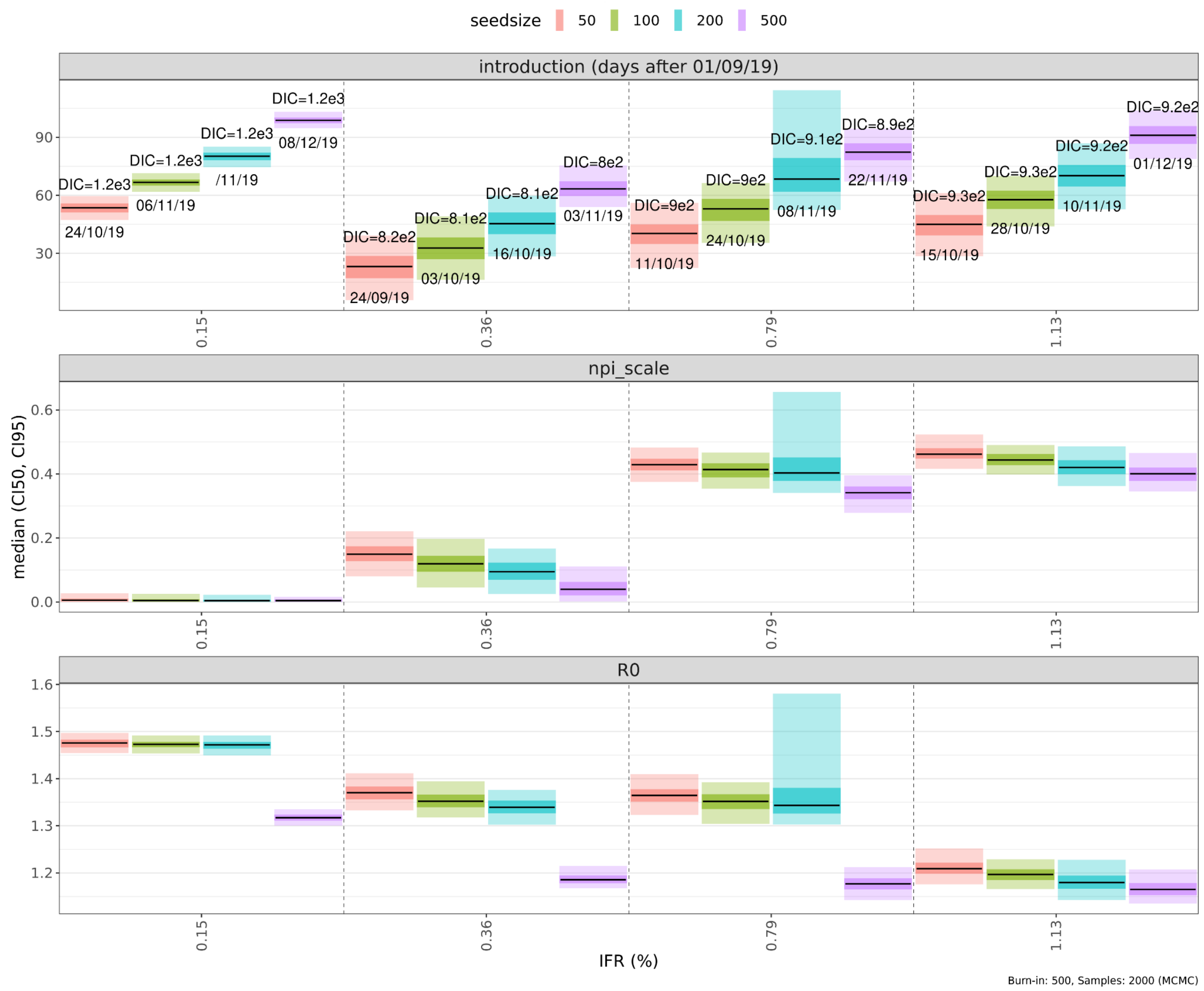


**SI Figure 15**. Posterior distributions of fitting parameters when importations are young adults between 20 and 30 years.
